# Describing a complex primary health care population in a learning health system to support future decision support and artificial intelligence initiatives

**DOI:** 10.1101/2022.03.01.22271714

**Authors:** Jacqueline K. Kueper, Jennifer Rayner, Merrick Zwarenstein, Daniel J. Lizotte

## Abstract

**Introduction:** Learning health systems (LHS) use data to improve care. Descriptive epidemiology to reveal health states and needs of the LHS population is essential for informing LHS initiatives, including development of decision support tools. To properly characterize complex populations, both simple statistical and artificial intelligence techniques can be useful. We present the first large-scale description of the population served by one of the first primary care LHS in North America.

**Objectives:** Our objective is to describe sociodemographic, clinical, and health care use characteristics of adult primary care clients served by the Alliance for Healthier Communities, which provides team-based primary health care through Community Health Centres (CHCs) across Ontario, Canada.

**Methods:** Using electronic health record data from 2009-2019 for all CHCs, we perform table-based summaries for each characteristic; and apply unsupervised leaning techniques to explore patterns of common condition co-occurrence, care provider teams, and care frequency.

**Results:** Of the 221,047 eligible clients, those at CHCs that primarily serve those most at risk (homeless, mental health, addictions) tend to have more chronic conditions and social determinants of health, which are also prominent in clients with multimorbidity. Most care is provided by physician and nursing providers, with heterogeneous combinations of other provider types. A subset of clients have many issues addressed within single-visits and there is within- and between-client variability in care frequency. Example methodological considerations learned for future LHS initiatives include the need to carefully consider the level of analysis and associated implications for data quality and target population, heterogeneity in conditions and care characteristics, and non-uniform risk profiles across the care history.

**Conclusions:** We demonstrate the use of methods from statistics and artificial intelligence, applied with an epidemiological lens, to provide an overview of a complex primary care population. In addition to substantive findings, we discuss implications for future LHS initiatives.

## Introduction

The recognized potential for analysis of electronic health record (EHR) data to inform health care delivery led to the formalization of the concept of a Learning Health System (LHS) in 2007: a socio-technical system characterized by iterative cycles of data-to-knowledge-to-practice feedback [1, 2]. LHS initiatives target quality improvement, research, or decision support; and usually rely on EHR data from the same population that the findings or end-product are intended to benefit [2–5]. These initiatives can support populations who have historically been excluded from medical research and clinical guideline development, such as those with complex health needs or barriers to participation [6–9].

Primary care, first contact care provided in a community setting over the life course, is inherently complex [10, 11]. The Alliance for Healthier Communities provides team-based primary health care through 72 Community Health Centres (CHCs) across Ontario to clients who face barriers to care and challenges, such as poverty and mental illness, that increase their risk for poor health [12–14]. Population health is a central element of their care model, and the Alliance officially adopted a LHS model in October 2020 [15, 16], making them one of few documented primary care LHSs in North America [5].

A LHS may pursue multiple initiatives to inform and improve care delivery. A first step towards any initiative is identifying needs of clients and providers, which is often driven by internal stakeholders [4]. Descriptive epidemiology is instrumental in outlining health states and needs of populations [17], and may be beneficial to add into these early stages of LHS development both to identify new areas to explore and to support existing ideas. For example, describing how clients are represented in EHR data at a population level may complement clinical experience to identify potential bias or misrepresentation that analyses need to account for to obtain meaningful results [18–20]. In addition to proposed LHS benefits, descriptive studies can contribute towards closing the gap in understanding about the basic functions of primary care in general [21].

To properly understand complex EHR data, we propose using both simple statistical techniques traditionally used in descriptive epidemiology and more complex techniques from artificial intelligence, applied with an epidemiological lens. Simple techniques alone may provide an oversimplified or incorrect view of certain characteristics, which could lead to ineffective or harmful decisions later-on. So, in pursuing our primary purpose of better understanding care provided by the Alliance, we explore the suitability of a variety of techniques for epidemiology of a separate primary care system with its own EHR.

We present the first large-scale descriptive and exploratory study of ongoing primary care clients served by the Alliance using statistical and machine learning methodology. Our *objective* is to summarize sociodemographic, clinical, and health care use characteristics of this population. We use unsupervised learning techniques to identify patterns of multimorbidity, care provider teams, and care access frequency. Findings will provide a foundation for future Alliance LHS initiatives, including those related to their existing interest in using EHR data to segment populations and tailor care. In addition to substantive findings, this work more generally demonstrates the application of an epidemiological lens and use of a variety of methods from statistics and artificial intelligence to effectively describe a complex population and contribute to early stages of a LHS.

## Methods

### Study population and data source

We use a de-identified extract of the centralized, structured EHR database from all CHCs; clients have unique identifiers to allow tracking of care over time. Issues addressed during care are recorded using Electronic Nomenclature and Classification Of Disorders and Encounters for Family Medicine (ENCODE-FM) [22] and International Classification of Disease (ICD)-10 vocabularies [23]. Primary care EHRs represent an open cohort; Supplementary Appendix 1 (Figure S1) shows the cohort size along calendar- and observation-based time definitions. Clients eligible for inclusion were over 18 years old in 2009, indicated a CHC as their primary care provider, and had at least one encounter at a CHC in 2009 to 2019. Any additional eligibility for specific analyses is described as needed below. We follow RECORD reporting guidelines (Supplementary Appendix 2)[24].

### General analysis plan

Sociodemographic, clinical, and health care use characteristics are defined in Supplementary Appendix 3 (Table S1). Methods specific to each category are described below; we perform “table-based summaries” for all, whereby categorical variables are summarized by counts and percentages, and continuous variables by the range, median, mean, and standard deviation. Where specified, findings are stratified by client multimorbidity status (defined below) or CHC “urban at-risk” (UAR) status, which are CHCs located in major urban geographical areas and serve priority populations defined by homelessness and/or mental health and substance use challenges [25]. CHCs without UAR designation still focus on clients with barriers to care but may be in rural or urban settings and do not solely serve clients with the aforementioned complexities [25].

**Table 1:**
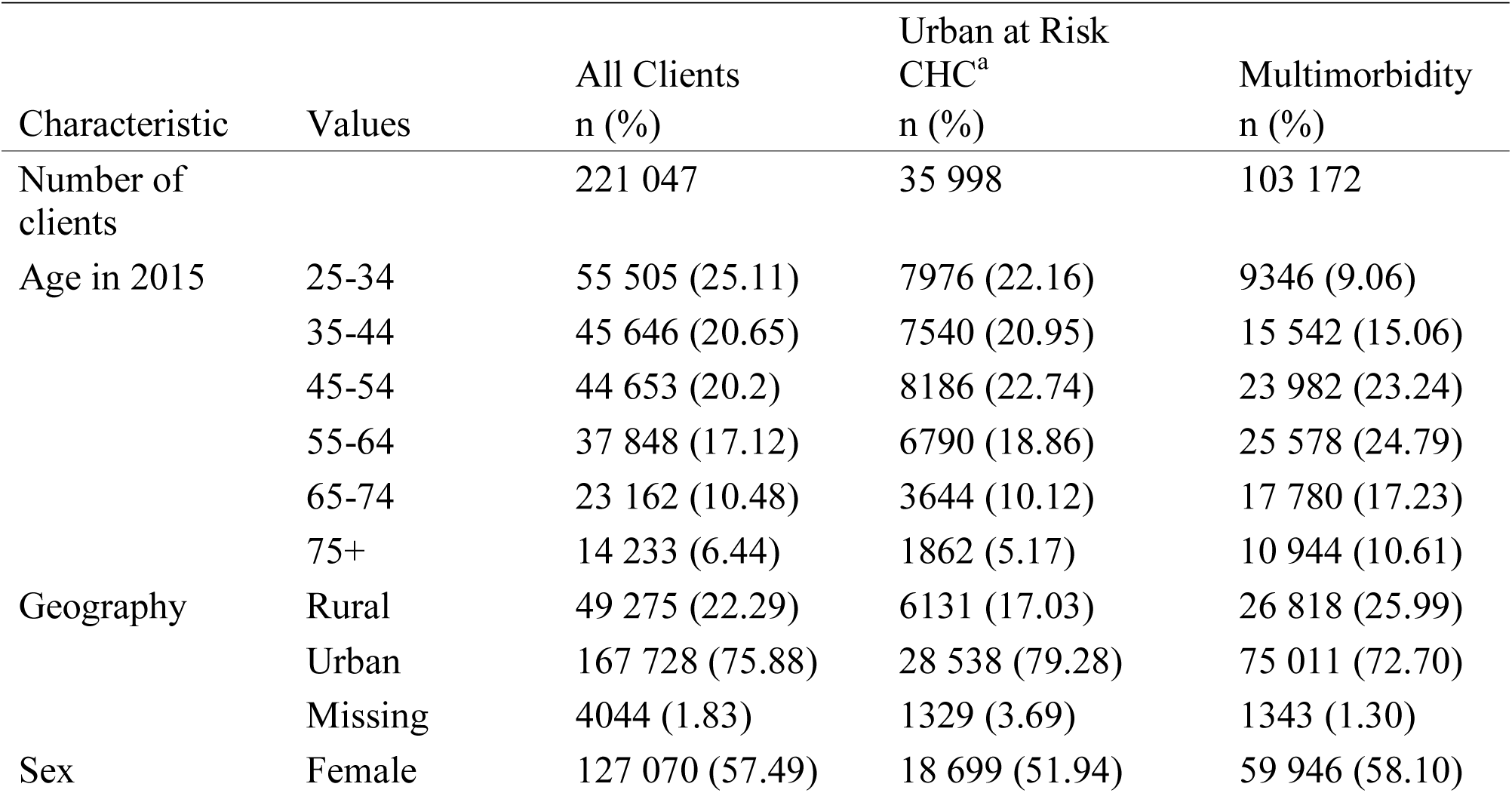
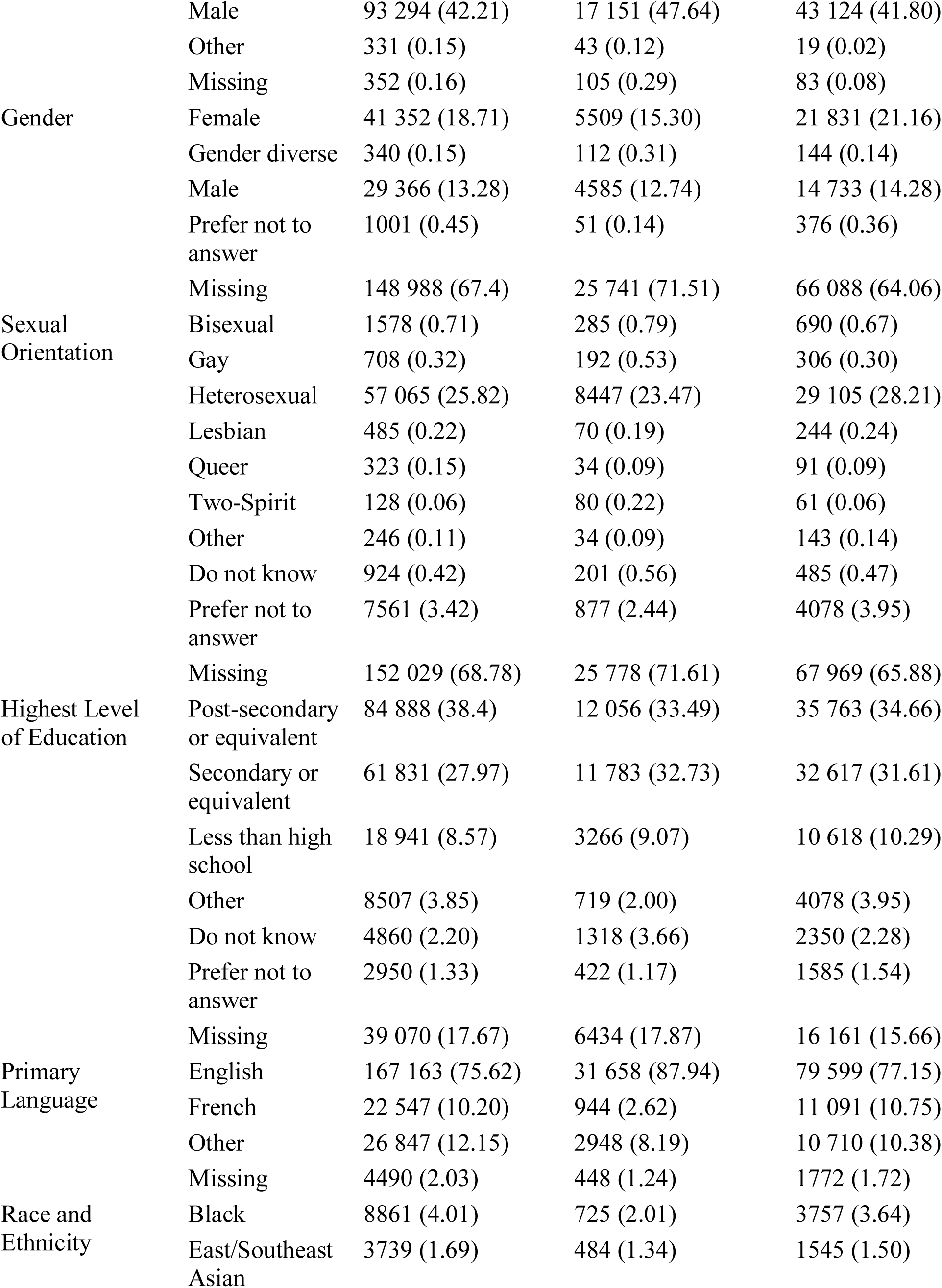
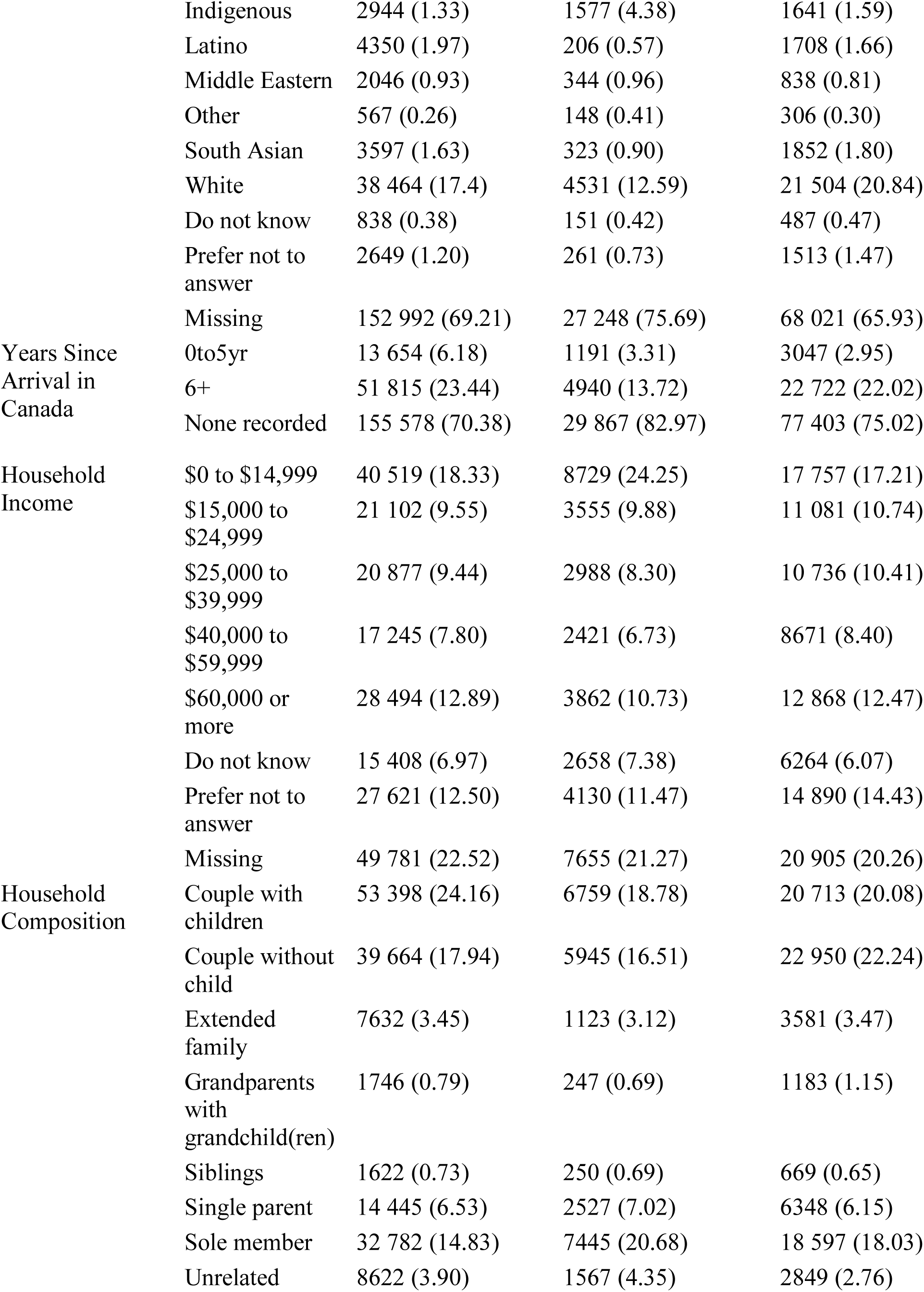
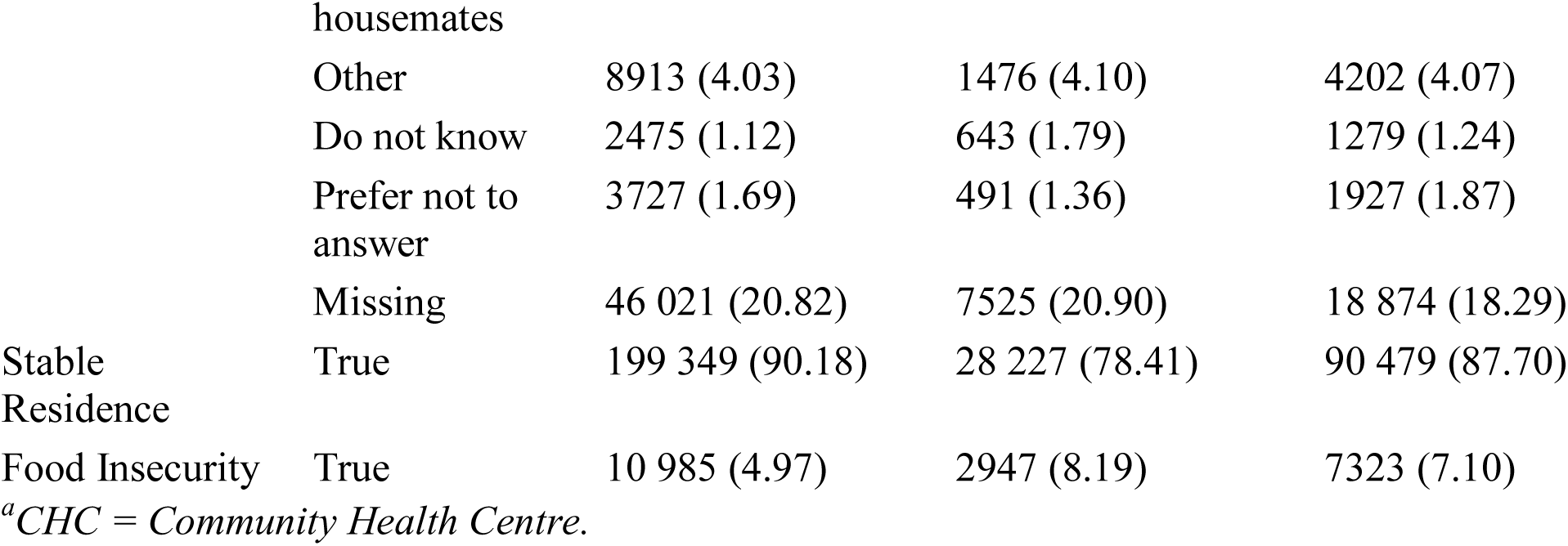
Sociodemographic characteristics.

### Sociodemographic characteristics

We provide table-based summaries for select fields from the structured EHR client characteristic table and certain ENCODE-FM-derived variables. Missingness of the former occurs at the 1) CHC or provider level, whereby a client is not asked about the characteristic and 2) client level, whereby a client is asked and preferred to not respond. Results are presented overall and stratified by UAR and multimorbidity status.

### Clinical characteristics

We describe 20 chronic conditions that define multimorbidity in PC research [26–28] and an additional four conditions of interest identified by Alliance stakeholders. For each condition, clients are assumed to receive related care upon the first record of a relevant code. Conditions are explored in single, composite, and pairwise manners.

#### Prevalence and incidence

To provide different perspectives on clinical complexity, we calculate two measures of prevalence and one measure of incidence for each of the 24 conditions. We also calculate prevalence of multimorbidity. Our primary multimorbidity definition, including for stratification, is presence of at least three of the 20 chronic conditions [26–28]. Multimorbidity of at least two conditions is also common and is presented for comparison [27].

(1) *Eleven-year period prevalence*, based on calendar time, to assess the burden of conditions over the entire observation period (2009-2019). For each condition, the number of clients who ever receive a condition indication is divided by an estimate of the average population size (technical details in Supplementary Appendix 3). Sensitivity analyses include the largest possible denominator: total number of eligible clients, and the smallest reasonable denominator: starting with the middle calendar year (2014), additional clients with at least one visit in adjacent years are added until no prevalence estimate is over 100%. Results are shown overall and UAR-stratified.
(2) *Observation-based period prevalence*, based on length of client observation, to assess the burden of conditions dependent on the number of years clients have received care at a CHC. To calculate this, clients are separated into 11 sub-cohorts based on the number of years (consecutive 365.25 day intervals, rounded up) between their first and last recorded events. For each sub-cohort and condition, the number of clients who ever receive a condition indication is divided by the number of clients in the sub-cohort. Results are presented as bar graphs.
(3) *Cumulative incidence*, to assess the rate of condition indications by days of observation. Cumulative incidence curves are plotted using the R package *survival* [29]. To prioritize capture of incident condition-related care, clients with conditions recorded in 2009 are excluded from this analysis.

#### Condition co-occurrence patterns

To assess co-occurrence for each pair of conditions while adjusting for all of the other conditions, we estimate an *Ising model* using R package *MRFcov* [30, 31] for all conditions except Hepatitis C (Alliance-suggested condition that overlaps with one of the 20 chronic conditions). We convert coefficients, representing the strength of association between each condition pair adjusted for all other conditions, to odds ratios and interpret size using Chen et al. (2010) guidelines [32]. We also view the top frequency-based co-occurrences.

### Health care use characteristics

We perform table-based summaries of provider and care access characteristics overall and stratified by UAR CHC, Rural Geography CHC, and client multimorbidity status.

#### Providers involved

To identify common care provider teams that clients are exposed to across their care histories, we use *non-negative matrix factorization (NMF)*[33] to identify frequently-occurring: 1) *“Ever-seen” teams* whereby dummy variables are used to indicate whether each provider type has ever been involved in care, and 2) *Relative “amount-seen” teams* based on volume of care whereby the number of events associated with each provider type are normalized within clients. For each version, analyses allowing 2,3,5,10, and 15 topics (provider teams) are run with the Python package *sklearn.decomposition.NMF* and the kullback-Leibler divergence distance metric [34]. Resulting topics are interpreted manually. Provider types are maintained as recorded in the EHR except “Other,” “Unknown,” and “Undefined” are combined. We also summarize the top frequency-based provider types involved in care and referrals. Eligible clients require at least one provider type indication in their EHR.

#### Care access patterns

*Complexity of care* is measured as the number of events (distinct issues addressed or types of care received) per visit (calendar day of access) to a CHC. *Care frequency* is measured as the number of calendar days at least one event is recorded per year (365.25 day intervals) and per quarter-year (90.30 day intervals). To investigate frequency of care in terms of magnitude and shape (changes in magnitude across care histories), we perform *time series clustering* with the K Medoids algorithm and dynamic time warping distance metric [35] for 1) *short-term clients* with 2-3 observation years and 2) *long-term clients* with 8-10 observation years. For each time interval and cohort, R package *dtwclust* [36] is used to identify 2,3,4, and 5 clusters. Performance is assessed using the silhouette score and visual inspection.

## Results

There are 221 047 eligible clients (Supplementary Appendix 3), of whom 64 504 (29.18%) received care at least once in 2009, 141 627 (64.07%) in 2019, and 40 704 (18.4%) received care in both years.

### Sociodemographic characteristics

Sociodemographic characteristics are described in **Table 1**, with remaining sub-strata in Supplementary Appendix 3 Table S2. The UAR CHCs tend to provide care to clients who are more commonly male, English-speaking, and have lower levels of education, household income, immigration, stable housing, and/or food security. Clients with multimorbidity tend to be older and more commonly female, reside in rural locations, and have lower levels of education, immigration, stable residence, and/or food security.

**Table 2:**
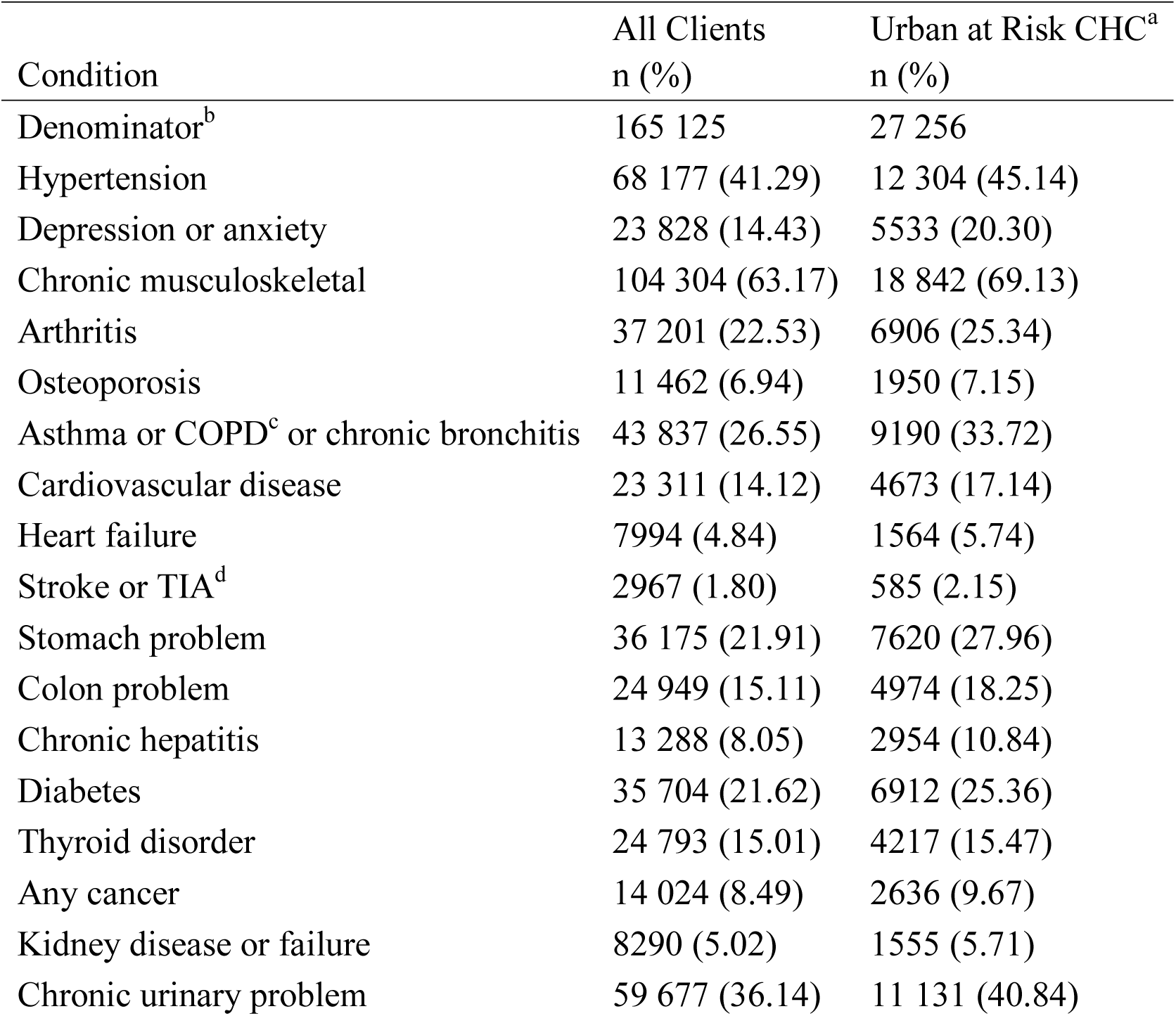
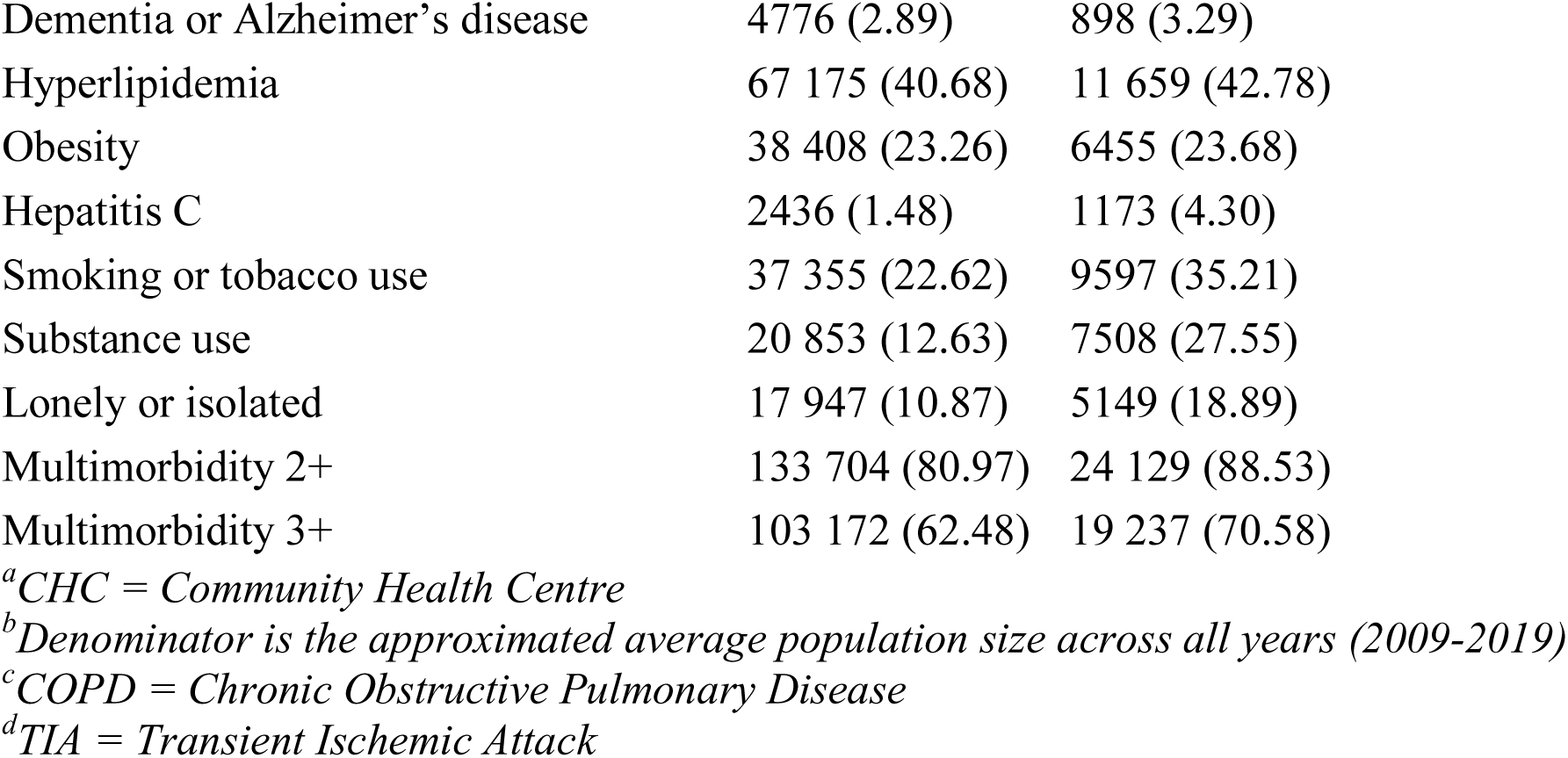
Eleven-year period prevalence

### Clinical characteristics

#### Prevalence and incidence

*Eleven-year period prevalence* estimates range from 1.48% (Hepatitis C) to 80.97% (multimorbidity of two conditions) overall, with generally higher estimates in UAR strata (**Table 2**). The low sensitivity estimate for the denominator is based on 2012-2015 (n=148 595).

*Observation-based period prevalence* estimates tend to increase with length of observation; however, cumulative incidence plots for the 156 543 (70.82%) clients without care recorded in 2009 show the rate of condition indications notably decreases after the first year of observation. Sample plots are in **Figure 1**; all are in Supplementary Appendix 1 (Figure S2 and S3).

**Figure 1:**
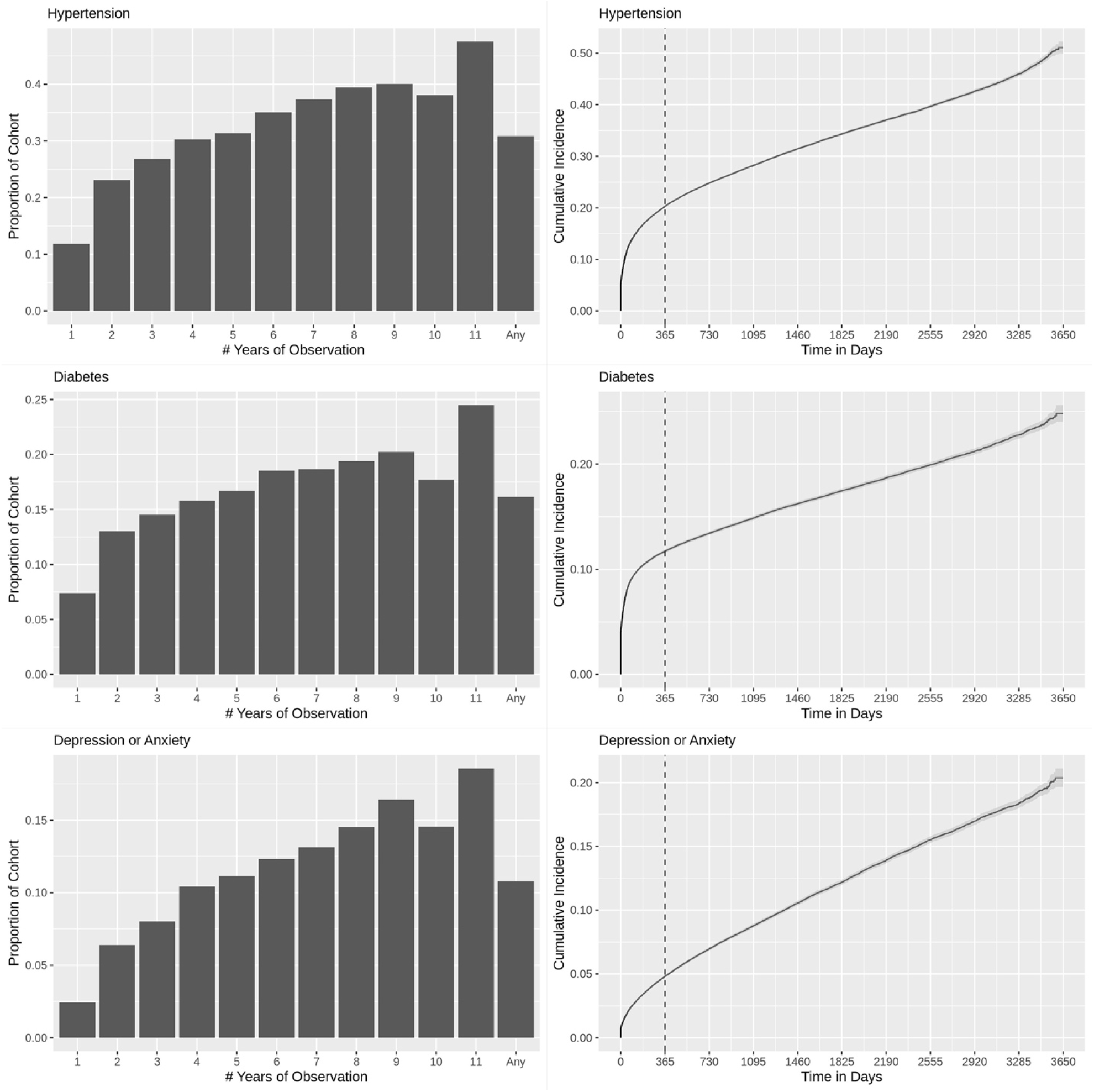
Example observation-based period prevalence and cumulative incidence plots. Left column: Observation-based period prevalence. Right column: Cumulative incidence by days of observation.

#### Condition co-occurrence patterns

Among the 103 172 (46.7%) clients with multimobidity of at least three chronic conditions, there are 25 162 unique combinations ranging in frequency from 1 (<0.1%) to 845 (0.4%) clients. **Figure 2** presents the *Ising model* results. Pairwise associations between conditions on the log-odds scale range from -0.82 (Osteoporosis—Obesity) to 2.93 (Kidney disease or failure— Chronic urinary problem). There are 1 large, 5 medium, 40 small, and 207 very small associations based on odds ratio magnitude. The five largest positive associations are 1) Kidney Disease or Failure—Chronic Urinary Problem, 2) Smoking or Tobacco Use—Substance Use, 3) Cardiovascular Disease—Heart Failure, 4) Hypertension—Hyperlipidemia, and 5)

**Figure 2:**
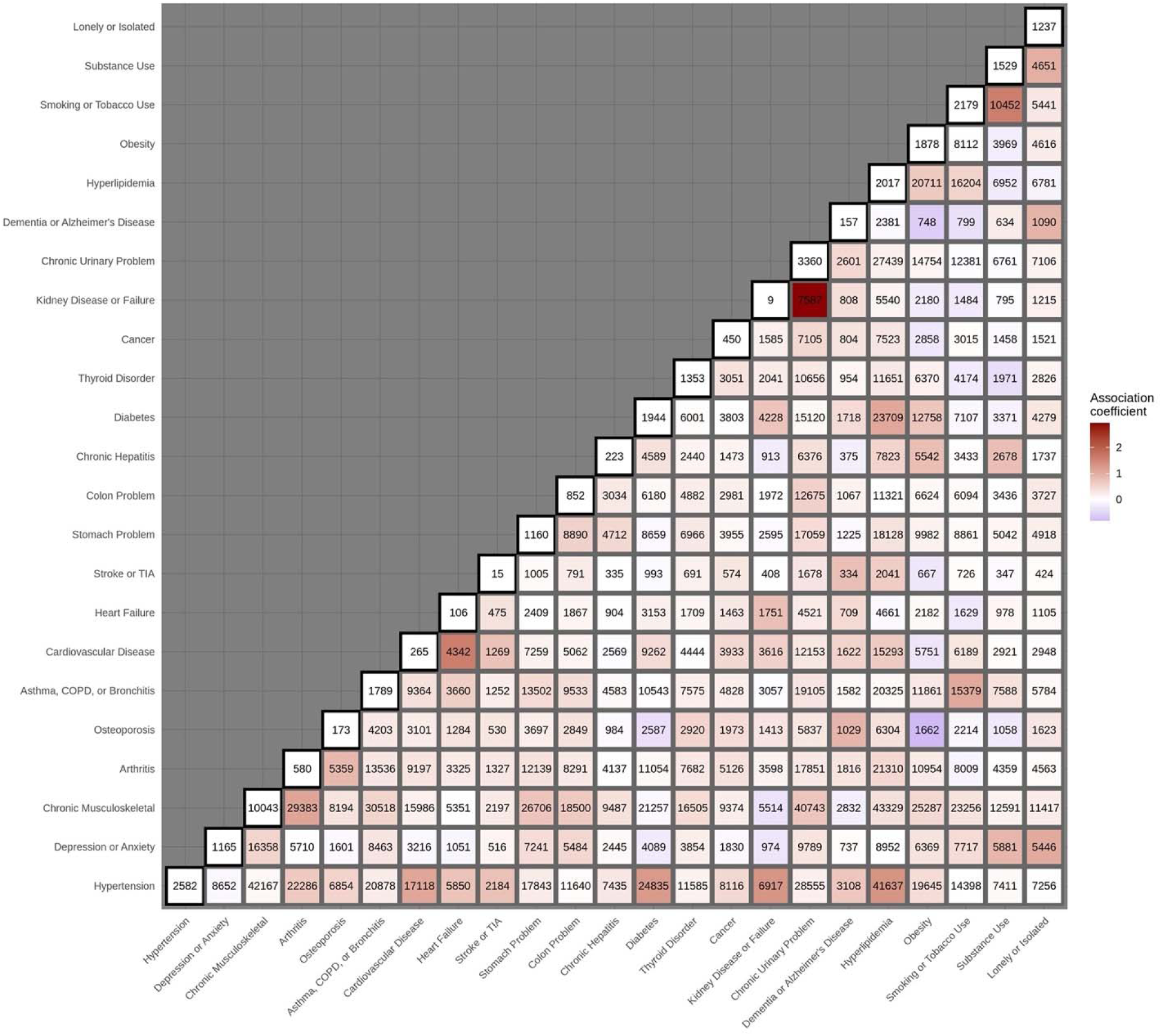
Condition co-occurrence patterns. Heatmap representing the results of the Ising model. Shading is relative to the edge weights or strength of condition co-occurrence. The numbers indicate raw counts in the data; diagonal counts represent clients who only have that single condition. *Legend*: TIA = Transient Ischemic Attack; COPD = Chronic Obstructive Pulmonary Disease.

Hypertension—Kidney Disease or Failure. In contrast, the top 5 co-occurring conditions based on raw frequency are 1) Hyperlipidemia—Chronic Musculoskeletal, 2) Hypertension—Chronic Musculoskeletal, 3) Hyperlipidemia—Hypertension, 4) Chronic Urinary Problem—Chronic Musculoskeletal, 5) Asthma or COPD or Chronic Bronchitis—Chronic Musculoskeletal. These directly correspond to the conditions with the highest marginal frequencies.

### Health care use characteristics

Table-based summaries of health care use characteristics are in Supplementary Appendix 3 (Table S3). In general, UAR and multimorbidity strata had higher health care use while rural geography CHCs were closer to the overall population.

#### Providers involved

There are 19 394 unique combinations of the 68 distinct provider types seen across the 220 806 (99.9%) clients with at least one provider type recorded. In terms of referrals, 102 088 (46.2%) clients had at least one internal and 143 922 (65.1%) had at least one external referral recorded. Note internal referrals may not capture “hallway referrals,” whereby a nearby provider provides a quick consult that is not formally recorded.

**Figure 3** shows results of the *NMF analysis*, listing the highest-weighted provider types in each topic down to a weight of 3. For the *ever-seen* provider team analysis, physician and nursing provider types emerged most prominently overall. In general, as the number of topics increases, additional provider types emerge and then split apart to dominate separate topics. Exceptions are the high-weighted pairings of nurse and physician and of registered practical nurse and nurse practitioner. Overall, 18 of the 68 possible provider types emerge prominently in at least one topic; only one (respirologist) does not also appear in the amount-seen analysis.

**Figure 3:**
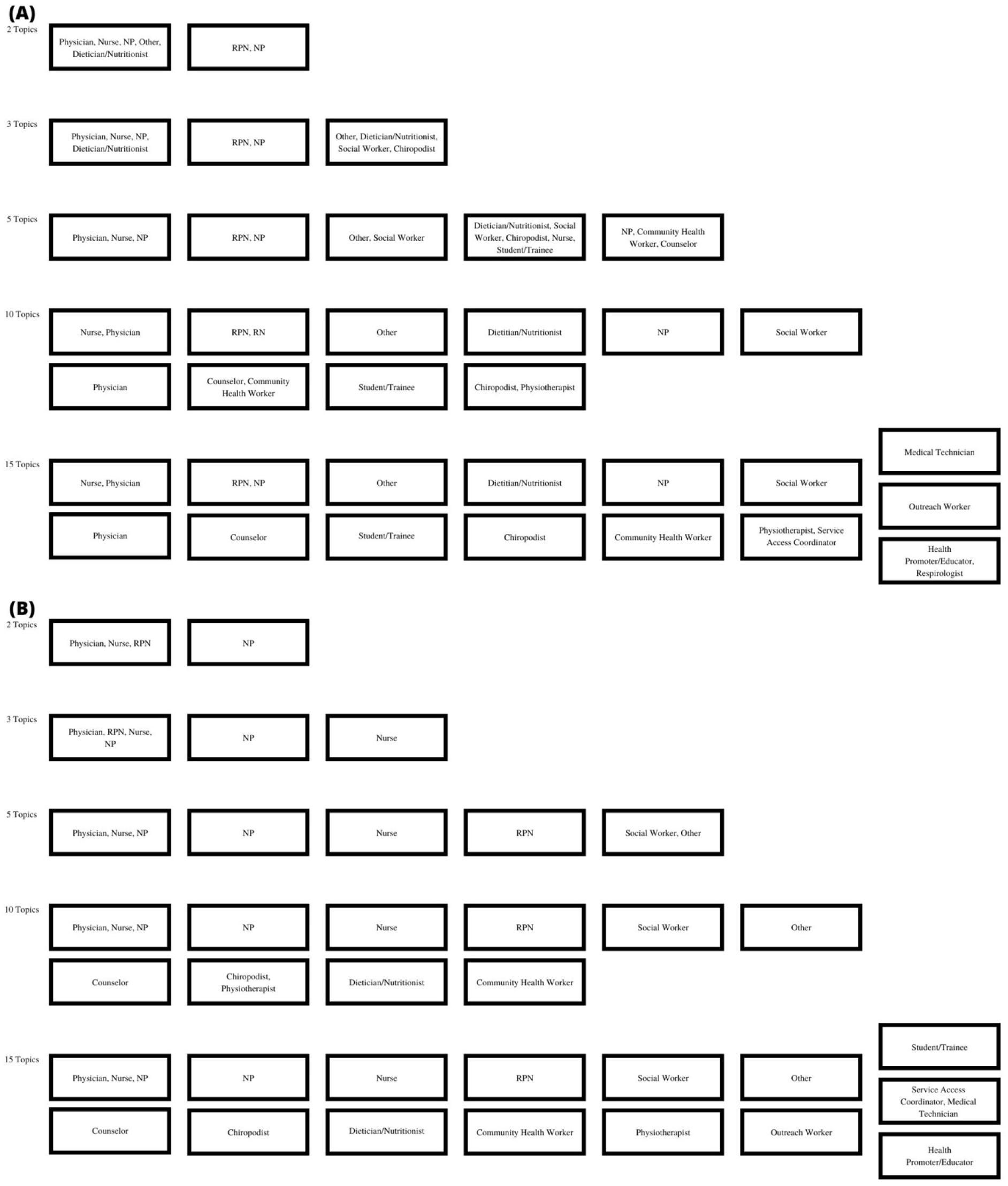
Common care provider teams. Boxes represent the topics resulting from the non-negative matrix factorization analysis for A) Ever-seen provider team analysis. B) Relative amount seen provider team analysis. Provider types are listed in order starting with the highest weighted provider; for any given topic, provider types with a weight less than three are not show. *Legend*: NP = Nurse Practitioner, RPN = Registered Practical Nurse.

The *amount-seen* provider team analysis has greater weight distributions between provider types within topics. For example, the first of the three-topic analysis has an approximate 1:1:1:6 ratio of care provided by nurse practitioner:nurse:registered practical nurse:physician. In both versions, about half of clients have a non-zero weight for only one of the first two topics; in the amount-seen analysis more clients remain non-zero weight on only only one topic as the number of topics increase, e.g. 16.6% versus 2.5% at five topics. In general, results suggests most clients receive the majority of care from physician, nurse practitioner, or nurse provider types, usually in combination with other provider types at a lower volume of care and with heterogeneous co-occurrence. An example of patterns that emerged for other provider types include differences in timing and weight of dietician/nutritionist and social worker providers between the two analyses. Interpreted alongside the most common provider and referrals types (Supplementary Appendix 3 (Table S4)), findings suggest referrals to dietitian/nutritionist are more common than to social worker, but frequent or longer-term care is more commonly provided by social workers.

#### Care access patterns

*Complexity of care* from a CHC-perspective is primarily low with 80.4% of client-visits associated with a single-issue and under 1.0% having over five issues addressed (higher intensity); however, from a client-perspective, 24 204 (11.0%) experience at least one visit with over five issues while 38 533 (17.4%) experience a maximum of one issue per visit across their care history. The mean *care access frequency* is 6 days per year (standard deviation=7.4). While 29 191 (13.2%) clients experience at least one year with over 25 days, 7455 (3.4%) average over 25 days per year across their entire care history. There are 8700 (3.94%) clients with at least one frequent care period (year with over 25 days care accessed) and complex care episode (visit with over 5 issues addressed).

For the *time series clustering* analyses, the short-term cohort includes 37 920 clients and 93 625 client-years of observation; the long-term cohort includes 42 855 clients and 387 035 client-years of observation. The silhouette score was always highest for two clusters (Supplementary Appendix 3 (Table S5)). Visual inspection of plots (**Figure 4**) shows high variability within and between clients.

**Figure 4:**
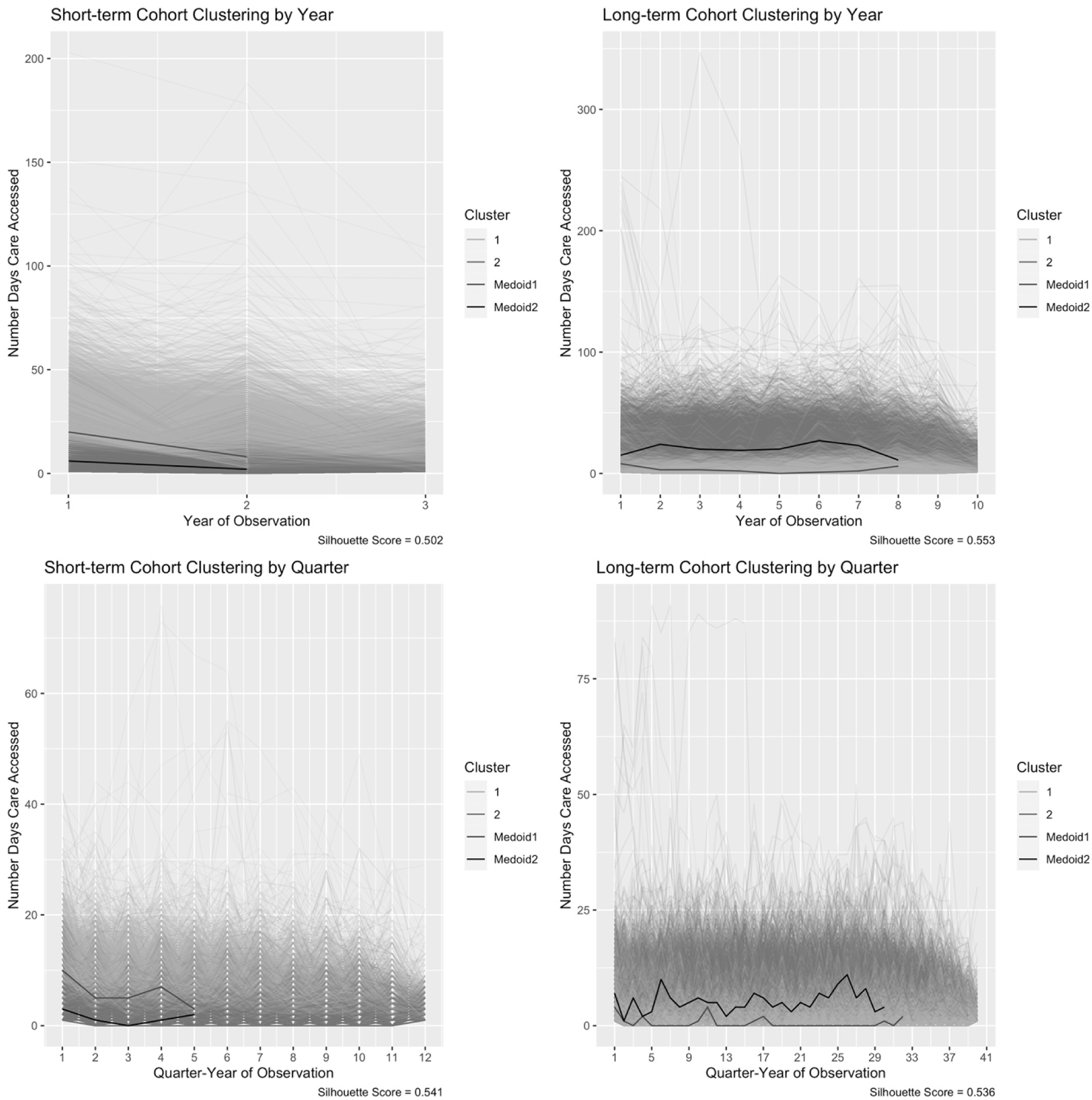
Care frequency clusters. Results from the four time series clustering analyses for each cohort and data-representation combination. Medoids are shown with raw time series data, separated by cluster number, for the number of clusters that resulted in the highest silhouette score (SS).

## Discussion

We used statistical and artificial intelligence techniques to summarize sociodemographic, clinical, and health care use characteristics captured in the EHRs of ongoing PC clients served by the Alliance. Substantive findings can motivate new topics for future LHS initiatives, or help to refine existing ideas and selection of performance measures for long-term evaluation of implemented interventions. Methods-related findings may inform the approaches used in these endeavours. While our discussion focuses on LHS initiatives, as with any epidemiological study, substantive results may be immediately useful to the population of interest, e.g., to inform clinic-level case management and onboarding of new clients.

### Sociodemographic characteristics

The CHC EHRs contain rich sociodemographic information, both the presence and absence of which is informative. Social determinants of health were more prevalent in UAR CHC and multimorbidity strata, and there appears to be evidence for the healthy immigrant effect [37]. Completeness rates vary by characteristic and may be due to client, provider, or CHC level decisions. For example, of the 72 059 (32.60%) clients asked about gender only 1001 (1.39%) preferred to not answer. In contrast, more clients, 171 266 (77.48%), were asked about household income but there was a higher tendency to not answer, 27 621 (16.13%). These findings align with a framework to assess selection bias in EHR data that suggests multiple mechanisms are usually responsible for missingness so the focus should be on “what data are observed [instead of missing] and why?”[38] While provider-level decisions may be due to inferring certain characteristics or prioritizing information needed for them to direct care, completeness rates are important for decision support tool performance, which can improve with social determinants of health information [39, 40].

When assessing data quality and completeness, which is emphasized by machine learning for EHR guidelines [2, 4, 19, 41, 42], the implications of pursuing LHS initiatives at different levels should also be considered. For example, a subset of CHCs capture self-reported measures of health, which are valuable research outcomes [43]. While these measures are not suitable for population level analyses, they should be considered for initiatives specific to the collecting CHCs.

### Clinical characteristics

#### Prevalence and incidence

In operationalizing morbidity measures, the denominator must be defined with the intended end-goal in mind. The *eleven-year period prevalence* estimates relate to a CHC-based perspective and are useful for long-term system-level planning, while the *observation-based period prevalence* estimates are more aligned with a client-based perspective and absolute measure of risk. Another consideration is that just as ICD-10 or ENCODE-FM codes do not guarantee true condition presence, the absence of care does not verify absence of conditions [44]. For example, clients may not seek PC when they are healthy, hospitalized, or experiencing barriers to care.

The *cumulative incidence* plots demonstrate that “risk” of condition codes is highest in the first year of observation. Clinically this makes sense, as new clients may have a build-up of unmet care needs. Nonetheless, there are important takeaways for LHS initiatives that require cohort construction. For example, predictive models developed for decision support need to account for the almost qualitative change in risk related to being a new client. Although this care pattern is somewhat unique to PC settings, methods developed for related problems may be useful. For example, accounting for variable lengths of stay in intensive care unit EHRs [45], or handling cold-starts and sparse data for recommender systems [46].

#### Condition co-occurrence patterns

There is a high prevalence of multimorbidity, but with so many different multimorbidity “compositions” it is hard to see how to make use of the category of multimorbidity. The *Ising model* demonstrates how to go beyond frequency-based comparisons and identify relationships between conditions irrespective of others, but again, this presents as a long tail problem, with very few combinations that are very prominent. PC decision support tools will face the challenge of making recommendations on many different and possibly co-occurring conditions. The majority of existing decision support tools and clinical guidelines focus on a single condition at a time; new techniques for providing evidence-based guidelines or recommendations for these vast numbers of combinations are needed [47–50].

### Health care use characteristics

#### Providers involved

While care for ongoing PC clients is typically led by physicians or nurse practitioners, CHCs include many provider types and LHS initiatives may choose to focus on particular provider type(s). The *NMF analyses* more easily identify reliable patterns of commonly seen provider types and teams than manually sifting through extensive count-based tables. Another use for NMF is dimensionality reduction or data pre-processing, whereby data are summarized to reduce the number of variables that need to be included in an analysis [33]. For example, NMF-derived topics could be used as inputs to a predictive model instead of separate variables to represent each provider type or specific, manually selected combinations.

#### Care access patterns

*Complexity of care* from a CHC system-level perspective is primarily low intensity (few problems addressed per visit). The subset of clients who experience higher care complexity do not tend to also have high frequency of care. Sporadic visit patterns may be due to unstable living arrangements or demanding life responsibilities; when there is uncertainty about when a client will return, providers may pack together multiple types of care. The marginal distribution of *care frequency* is right-skewed without a distinct break; most clients experience lower care frequency, but higher frequencies are also observed. In contrast to expectations, we did not identify consistent, distinct client groupings through the time-series clustering, e.g., to indicate a subpopulation of “frequent visitors.” This may be due to restrictions in the types of similarity that dynamic time warping captures. Future analyses could try a different similarity metric or including covariates to account for baseline variability.

### Strengths and Limitations

Strengths include the deep interdisciplinary approach used to assess complex, longitudinal EHR data. We used chronic condition definitions recommended for PC research;[26–28] although the algorithms have not been validated for CHCs specifically. Our broad cohort definition supports a high-level overview of the population, but may not be appropriate for specific research questions.

### Conclusions

This study demonstrates the use of simple statistics and artificial intelligence techniques, applied with an epidemiological lens, to describe EHR data from a budding LHS. Substantive findings lay a foundation for future Alliance initiatives and may be informative for other organizations serving complex PC populations.

Key suggestions for future LHS initiatives include the need to carefully deliberate the level of analysis, or who a given initiative should be targeted at (e.g., population or specific CHCs, one or many clinical presentations, all or subset of providers), and the associated implications for how clients will be represented in the data. Representation will depend on analytical-, system-, provider-, and client-level factors. Decision support initiatives need to consider heterogeneity in conditions and care access patterns, including non-uniform risk of condition indications across observation history.

## Data Availability

The data underlying this article were securely accessed from the Alliance for Healthier Communities. The data cannot be shared publicly due to their sensitive nature, as agreed upon in the ethics agreement.

## Acknowledgements

This work was supported by the Canadian Institutes of Health Research Canadian Graduate Scholarship-Doctoral to JKK with supervisor DJL.

## Ethics statement

This study was approved by Western University Review Ethics Board project ID 111353.

## Supplementary appendices

*Appendix 1* includes extended results presented through figures.

*Appendix 2* includes the RECORD reporting guideline checklist.

*Appendix 3* includes extended results presented through tables and technical details.

## Abbreviations

CHC: Community Health Centre
EHR: Electronic Health Record
ENCODE-FM: Electronic Nomenclature and Classification Of Disorders and Encounters for Family Medicine
ICD-10: International Classification of Disease - Version 10
LHS: Learning Health System
NMF: non-negative matrix factorization
PC: Primary Care
UAR: Urban At-Risk

## Supplementary Material

## Appendix 1

**Figure S1:**
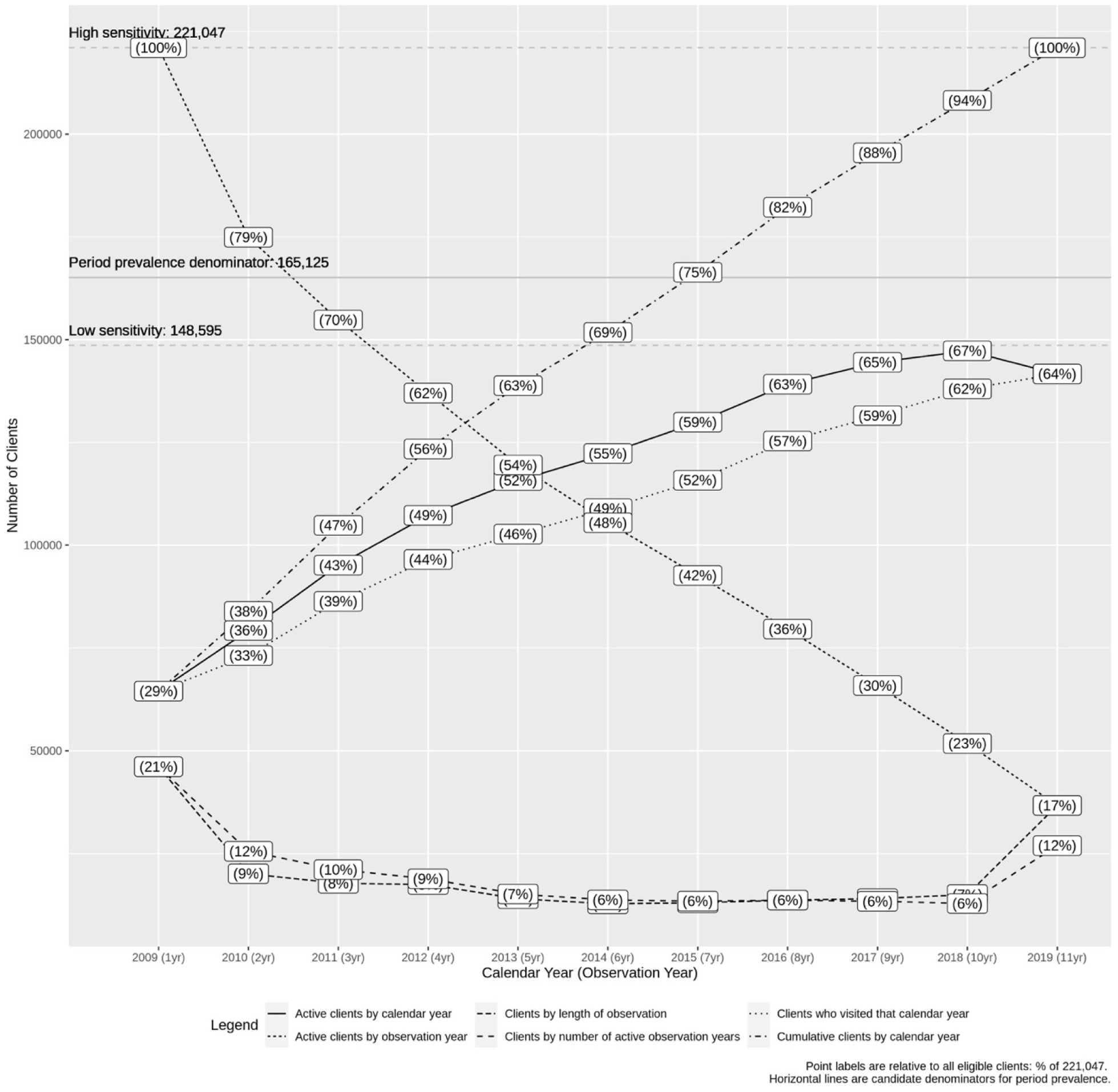
Cohort size by calendar- and observation-based time. Active clients have at least one event during or after the year (calendar- or observation-based) of interest (gap years counted). The number of active observation years refers to the number of 365.25 day periods, counted from the first calendar date that an event was recorded for that client, that clients have at least one event recorded (gap years not counted). Length of observation refers to the number of years from the first to the last year that at least one event is recorded during (gap years counted). Cumulative clients refers to the number of clients who have had at least one event during or before the year of interest. *Legend:* COPD = Chronic Obstructive Pulmonary Disease; TIA = Transient Ischemic Attack; AD = Alzheimer’s Disease.

**Figure S2:**
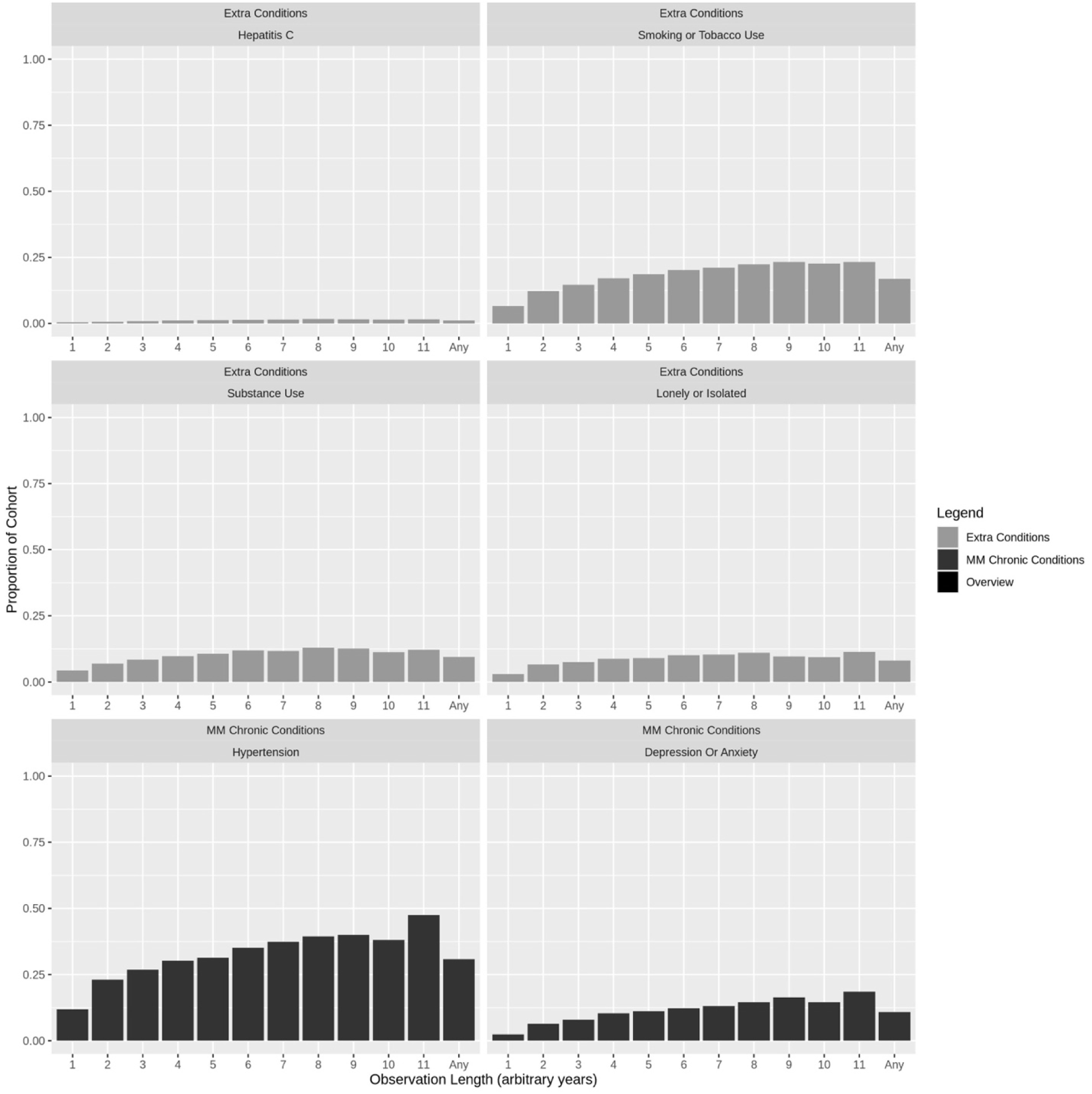
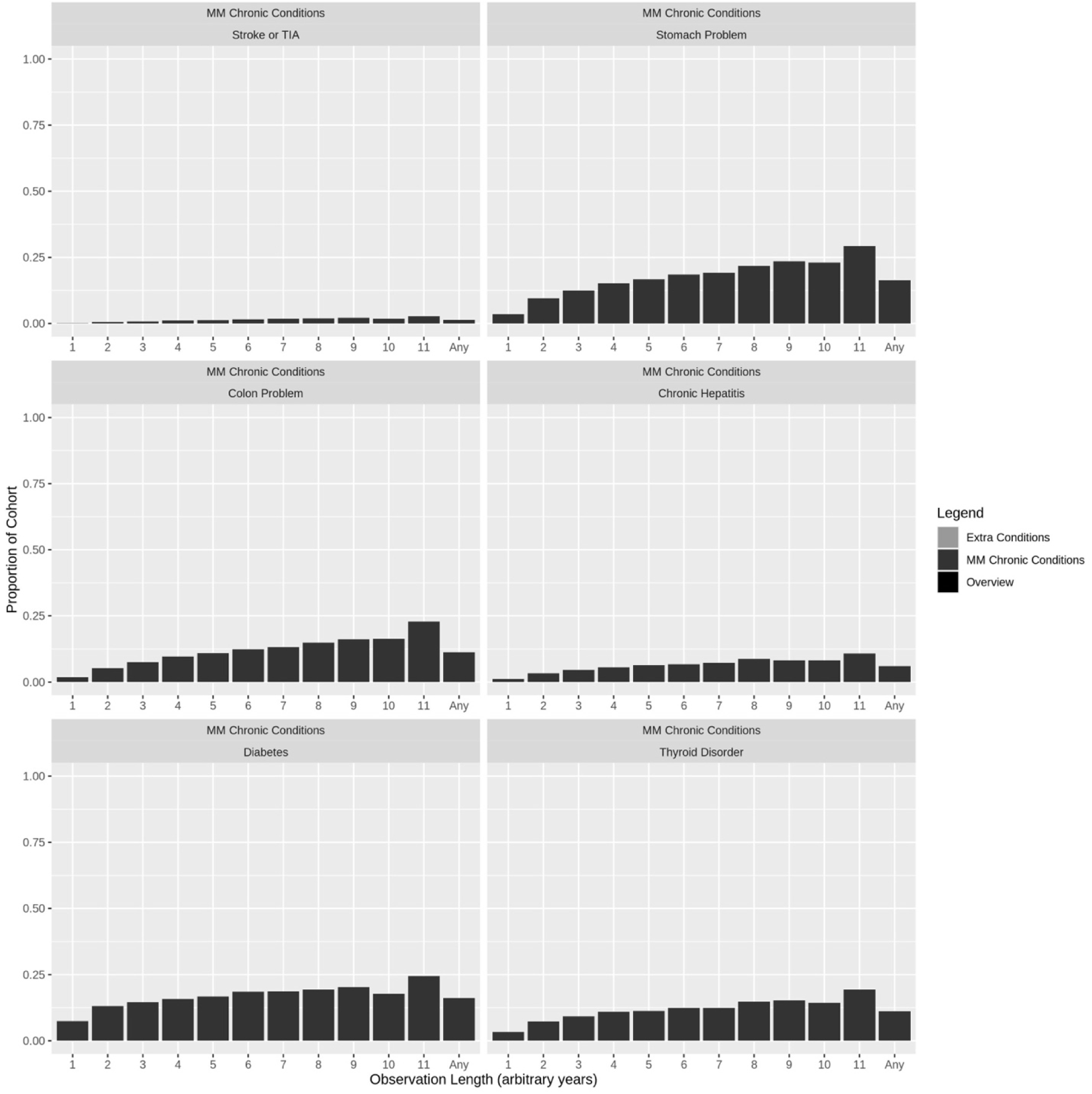
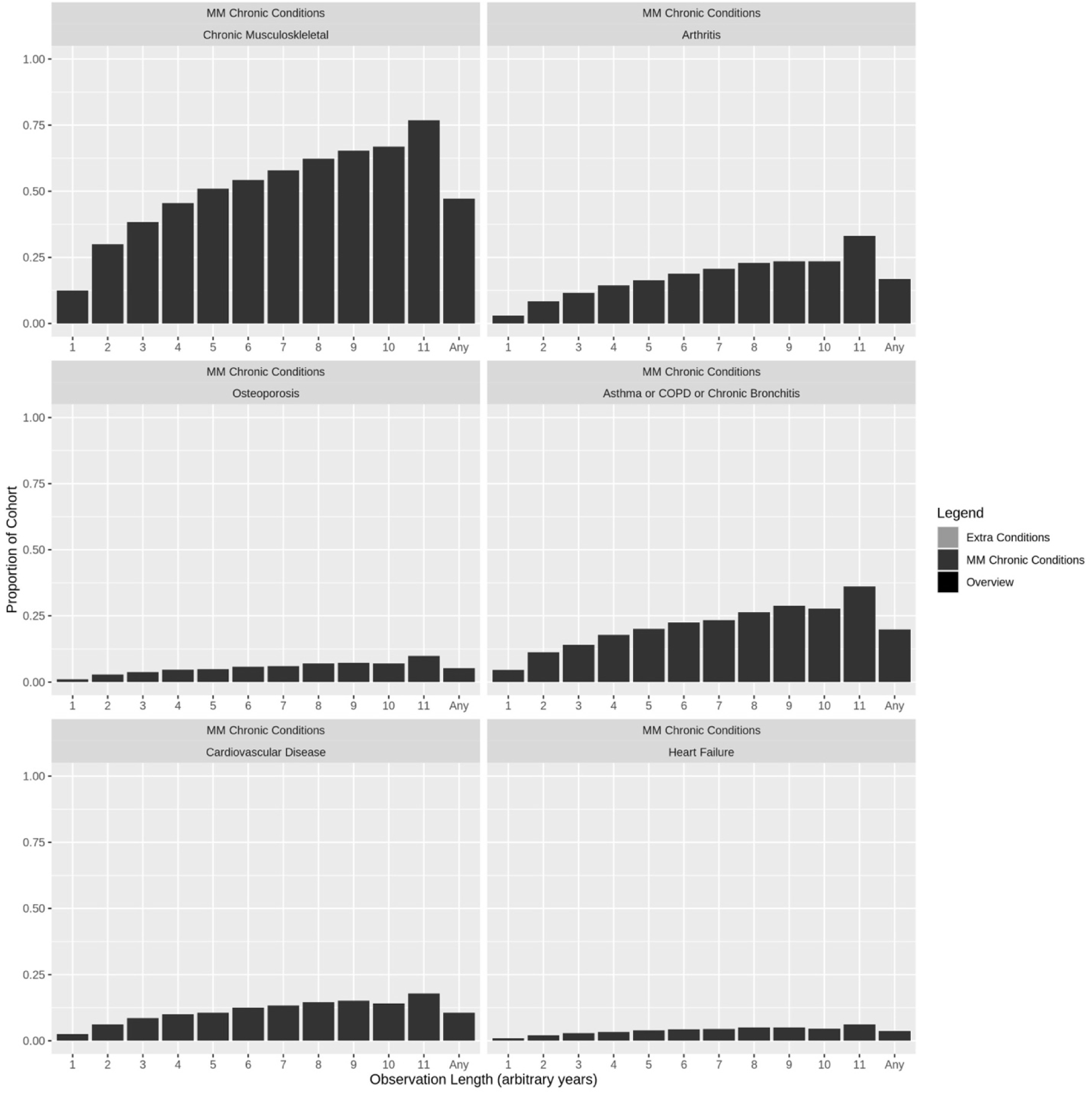
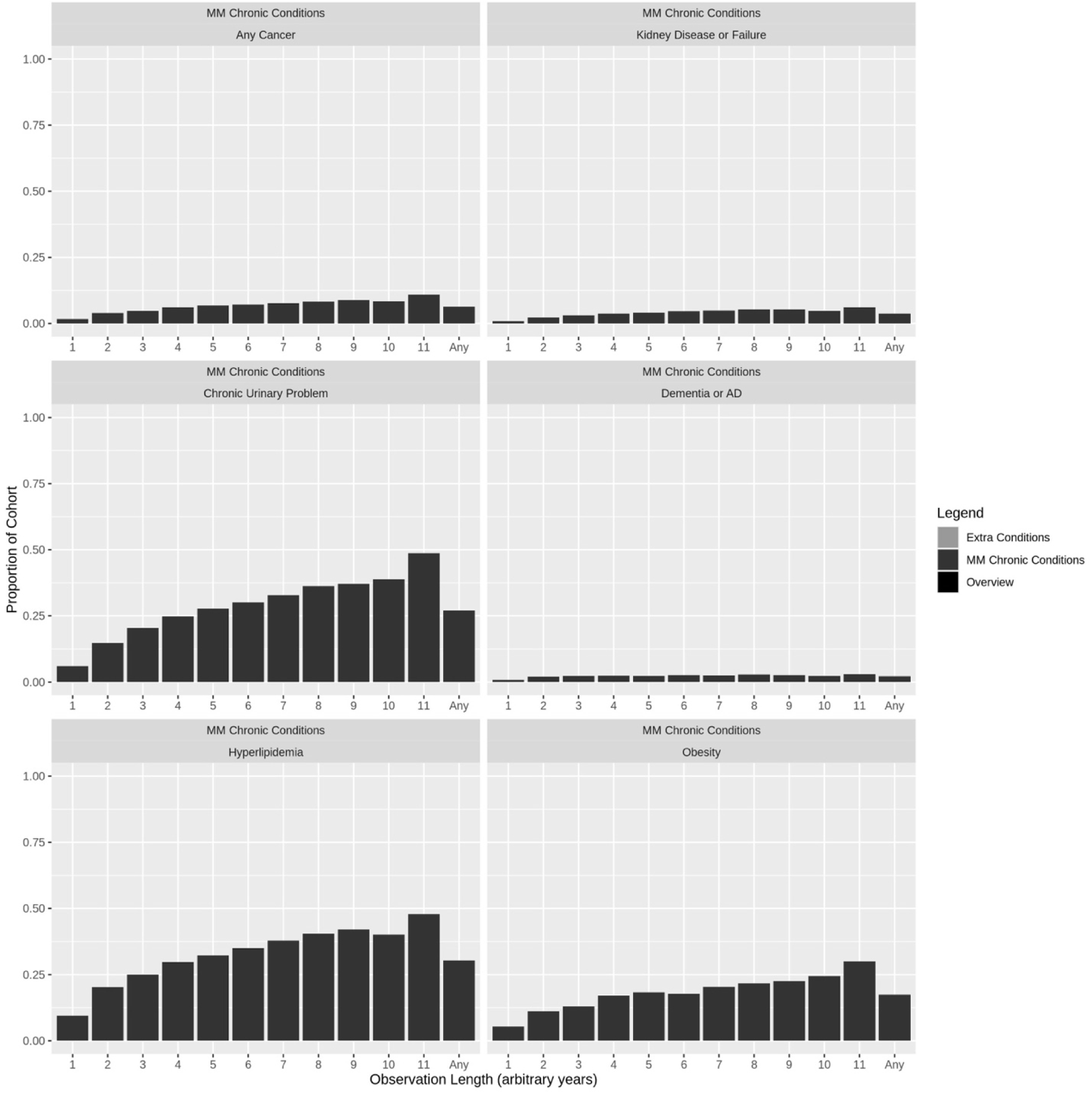
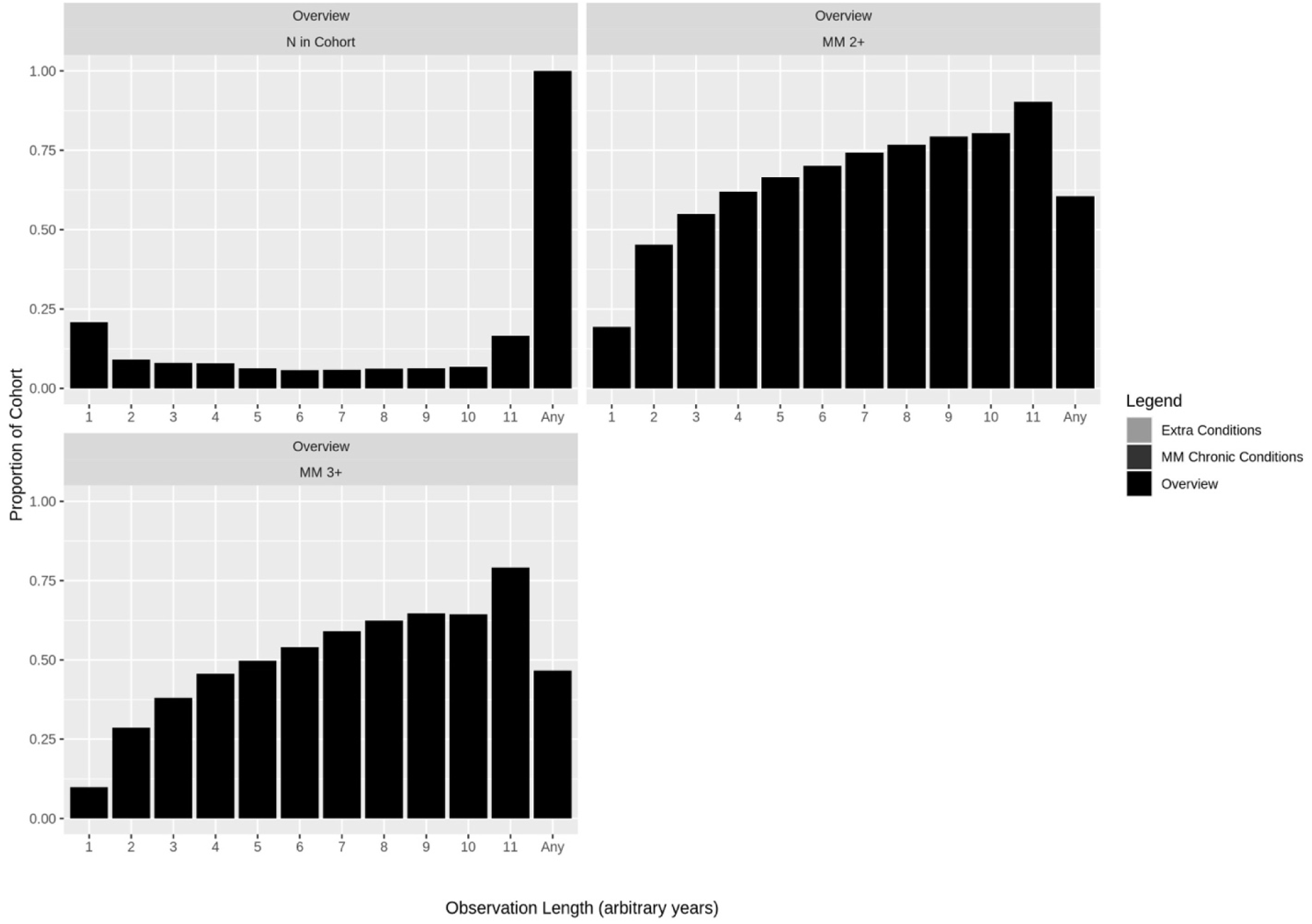
Observation-based period prevalence. Each bar represents the proportion of clients within that observation-based cohort (years are arbitrary 365.25 day consecutive periods between the first and last recorded events) that have at least one indication of the condition of interest across their entire observation history. Conditions are grouped to represent 1) Extra conditions of interest to Alliance stakeholders, 2) 20 chronic conditions, which make up multimorbidity (MM) status, and 3) Overview indicators for the cohorts. *Legend:* COPD = Chronic Obstructive Pulmonary Disease; TIA = Transient Ischemic Attack; AD = Alzheimer’s Disease.

**Figure S3:**
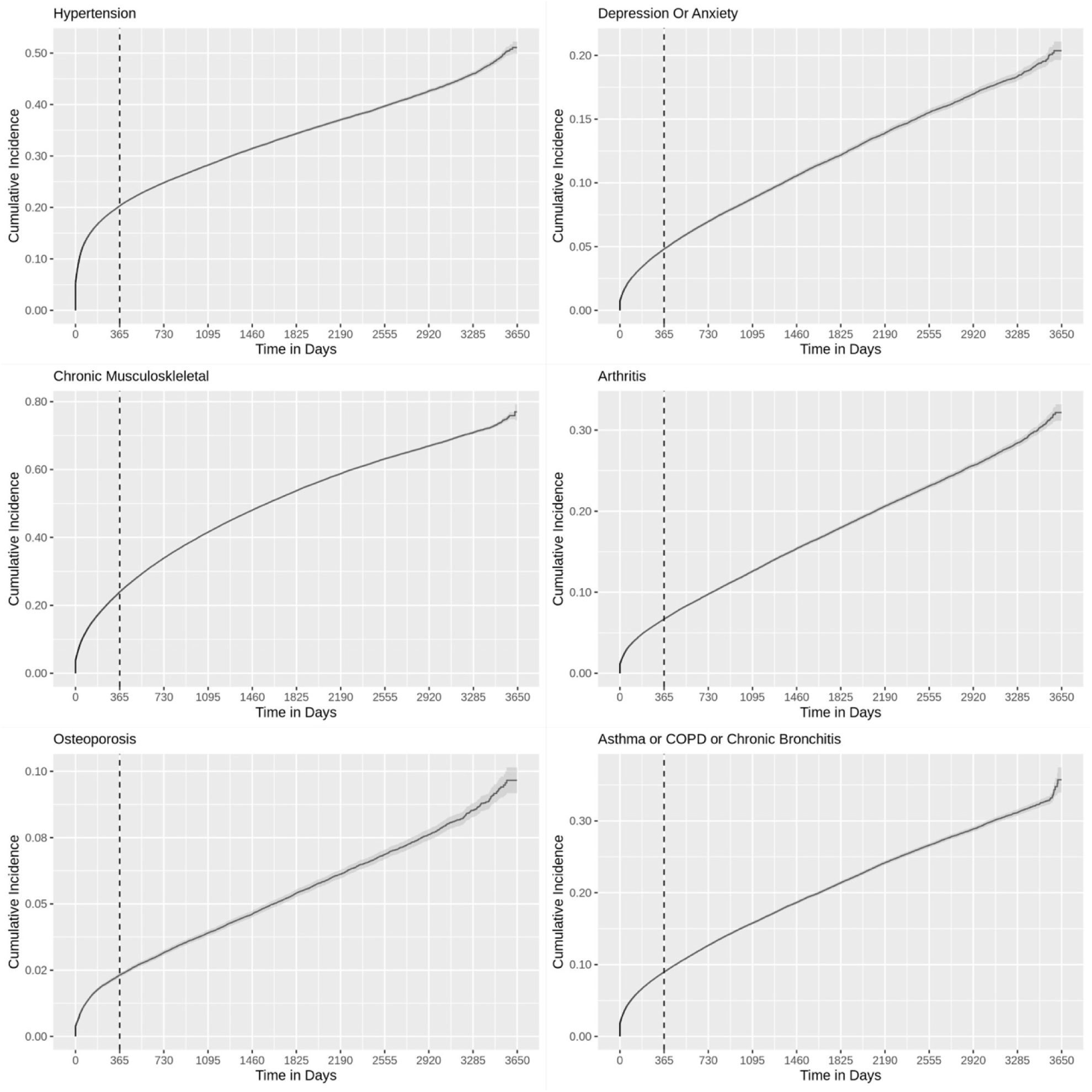
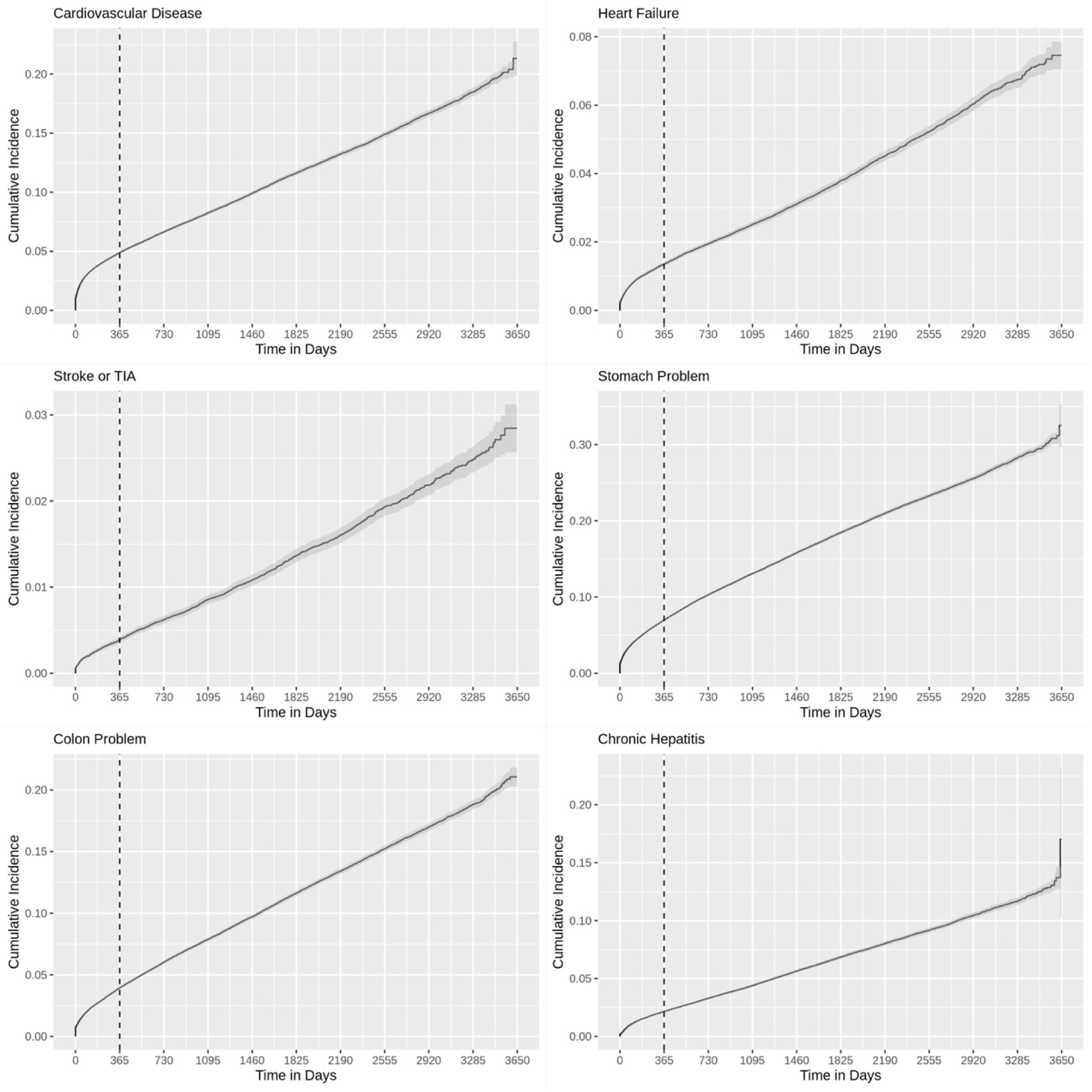
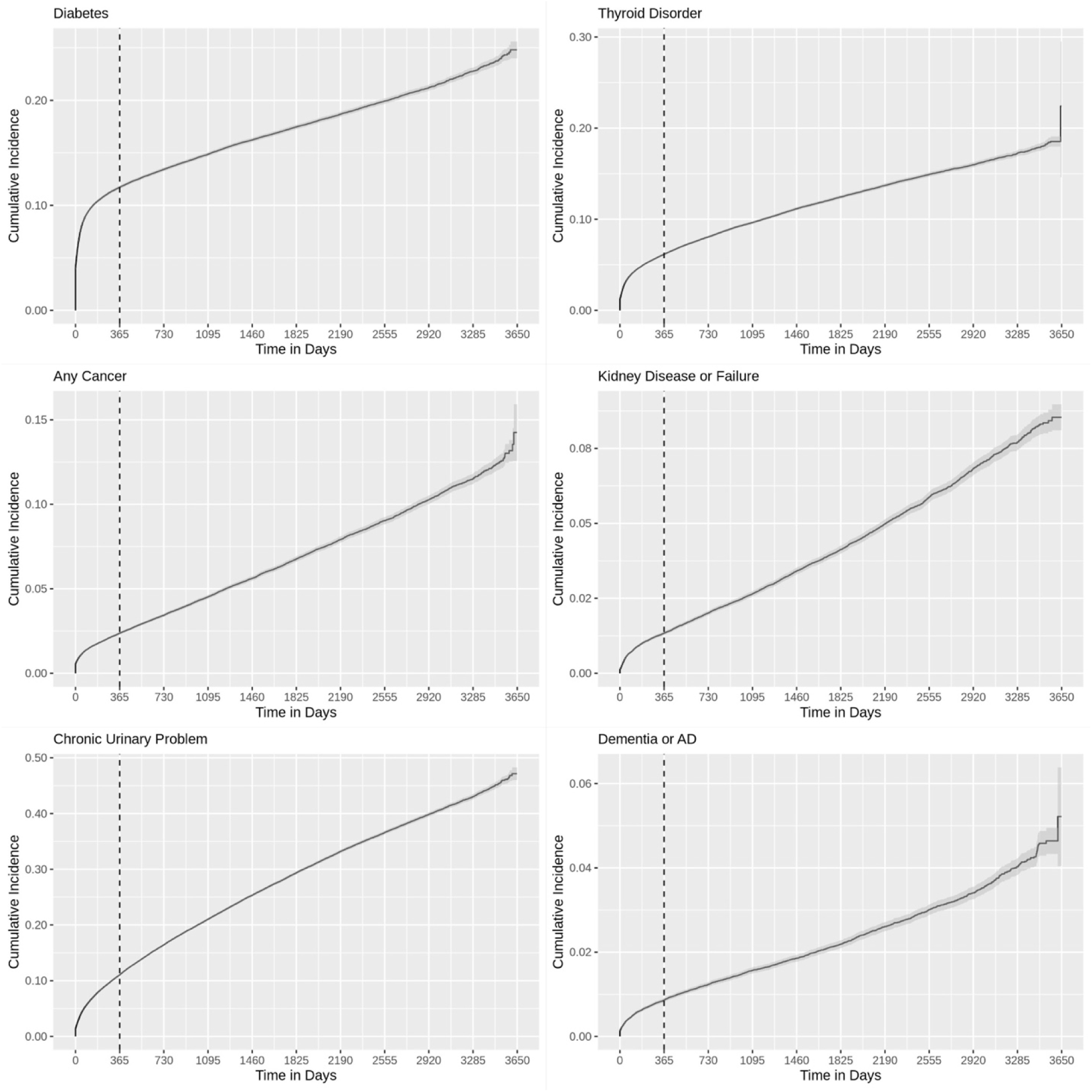
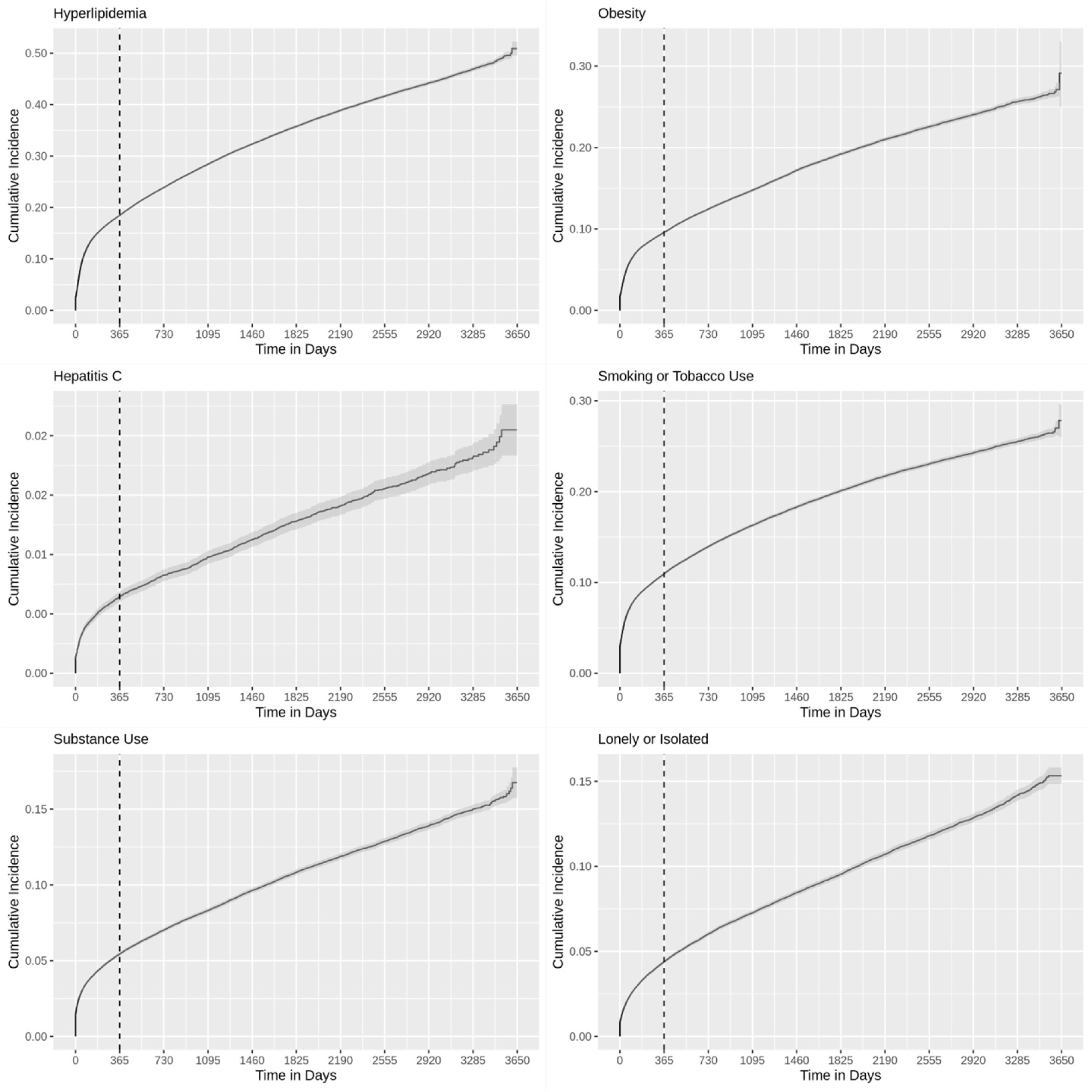
Cumulative incidence. Cumulative incidence plots by days of observation since the first recorded event. Clients eligible for this analysis must not have any care recorded in the first calendar-year of available data (2009).

## Appendix 2

**The RECORD statement – checklist of items, extended from the STROBE statement, that should be reported in observational studies using routinely collected health data.**

|  | Item No. | STROBE items | Location in manuscript where items are reported | RECORD items | Location in manuscript where items are reported |
| --- | --- | --- | --- | --- | --- |
| **Title and abstract** |  |  |  |  |  |
|  | 1 | (a) Indicate the study's design with a commonly used term in the title or the abstract (b) Provide in the abstract an informative and balanced summary of what was done and what was found | Abstract | RECORD 1.1: The type of data used should be specified in the title or abstract. When possible, the name of the databases used should be included.<br>RECORD 1.2: If applicable, the geographic region and timeframe within which the study took place should be reported in the title or abstract.<br>RECORD 1.3: If linkage between databases was conducted for the study, this should be clearly stated in the title or abstract. | Abstract |
| **Introduction** |  |  |  |  |  |
| Background rationale | 2 | Explain the scientific background and rationale for the investigation being reported | Introduction |  | Introduction |
| Objectives | 3 | State specific objectives, including any prespecified hypotheses | Introduction |  | Introduction |
| **Methods** |  |  |  |  |  |
| Study Design | 4 | Present key elements of study | Introduction, |  | Introduction, |
|  |  | design early in the paper | Methods – Study population and data source |  | Methods – Study population and data source |
| Setting | 5 | Describe the setting, locations, and relevant dates, including periods of recruitment, exposure, follow-up, and data collection | Methods – Study population and data source; additional details specific to analyses are presented under the appropriate sub-heading |  | Methods – Study population and data source; additional details specific to analyses are presented under the appropriate sub-heading |
| Participants | 6 | (a) *Cohort study* - Give the eligibility criteria, and the sources and methods of selection of participants. Describe methods of follow-up<br>*Case-control study* - Give the eligibility criteria, and the sources and methods of case ascertainment and control selection. Give the rationale for the choice of cases and controls<br>*Cross-sectional study* - Give the eligibility criteria, and the sources and methods of selection of participants<br>(b) *Cohort study* - For matched studies, give matching criteria and number of exposed and unexposed | Methods | RECORD 6.1: The methods of study population selection (such as codes or algorithms used to identify subjects) should be listed in detail. If this is not possible, an explanation should be provided.<br>RECORD 6.2: Any validation studies of the codes or algorithms used to select the population should be referenced. If validation was conducted for this study and not published elsewhere, detailed methods and results should be provided.<br>RECORD 6.3: If the study involved linkage of databases, consider use of a flow diagram or other graphical display to demonstrate the data linkage process, including the number of | Methods |
|  |  | *Case-control study* - For matched studies, give matching criteria and the number of controls per case |  | individuals with linked data at each stage. |  |
| Variables | 7 | Clearly define all outcomes, exposures, predictors, potential confounders, and effect modifiers. Give diagnostic criteria, if applicable. | Methods, Supplementary Table S1 | RECORD 7.1: A complete list of codes and algorithms used to classify exposures, outcomes, confounders, and effect modifiers should be provided. If these cannot be reported, an explanation should be provided. | Supplementary Table S1 |
| Data sources/<br>measurement | 8 | For each variable of interest, give sources of data and details of methods of assessment (measurement). Describe comparability of assessment methods if there is more than one group | Methods, Supplementary Table S1 |  | Methods, Supplementary Table S1 |
| Bias | 9 | Describe any efforts to address potential sources of bias | N/A |  | N/A |
| Study size | 10 | Explain how the study size was arrived at | Methods |  | Methods |
| Quantitative variables | 11 | Explain how quantitative variables were handled in the analyses. If applicable, describe which groupings were chosen, and why | Methods, Supplementary Table S1 |  | Methods, Supplementary Table S1 |
| Statistical methods | 12 | (a) Describe all statistical methods, including those used to control for confounding<br>(b) Describe any methods used to examine subgroups and interactions<br>(c) Explain how missing data<br>were addressed<br>(d) *Cohort study* - If applicable, explain how loss to follow-up was addressed<br>*Case-control study* - If applicable, explain how matching of cases and controls was addressed<br>*Cross-sectional study* - If applicable, describe analytical methods taking account of sampling strategy<br>(e) Describe any sensitivity analyses | Methods |  | Methods |
| Data access and cleaning methods |  | .. |  | RECORD 12.1: Authors should describe the extent to which the investigators had access to the database population used to create the study population.<br>RECORD 12.2: Authors should provide information on the data cleaning methods used in the study. | Methods, Supplementary Table S1 |
| Linkage |  | .. |  | RECORD 12.3: State whether the study included person-level, institutional-level, or other data linkage across two or more databases. The methods of linkage and methods of linkage quality evaluation should be provided. | No linkage |
| **Results** |  |  |  |  |  |
| Participants | 13 | (a) Report the numbers of individuals at each stage of the | Supplementary Appendix 3 | RECORD 13.1: Describe in detail the selection of the persons included in the | Methods |
|  |  | study (*e.g.*, numbers potentially eligible, examined for eligibility, confirmed eligible, included in the study, completing follow-up, and analysed)<br>(b) Give reasons for non-participation at each stage.<br>(c) Consider use of a flow diagram |  | study (*i.e.*, study population selection) including filtering based on data quality, data availability and linkage. The selection of included persons can be described in the text and/or by means of the study flow diagram. |  |
| Descriptive data | 14 | (a) Give characteristics of study participants (*e.g.*, demographic, clinical, social) and information on exposures and potential confounders<br>(b) Indicate the number of participants with missing data for each variable of interest<br>(c) *Cohort study* - summarise follow-up time (*e.g.*, average and total amount) | Results,<br>Supplementary<br>Appendix 3 |  |  |
| Outcome data | 15 | *Cohort study* - Report numbers of outcome events or summary measures over time<br>*Case-control study* - Report numbers in each exposure category, or summary measures of exposure<br>*Cross-sectional study* - Report numbers of outcome events or summary measures | Results,<br>Supplementary<br>Appendix 3 |  |  |
| Main results | 16 | (a) Give unadjusted estimates and, if applicable, confounder-adjusted estimates and their<br>precision (e.g., 95% confidence interval). Make clear which confounders were adjusted for and why they were included<br>(b) Report category boundaries when continuous variables were categorized<br>(c) If relevant, consider translating estimates of relative risk into absolute risk for a meaningful time period | Results,<br>Supplementary<br>Appendix 3 |  |  |
| Other analyses | 17 | Report other analyses done—e.g., analyses of subgroups and interactions, and sensitivity analyses | Results, Supplementary Appendix 1,3 |  |  |
| **Discussion** |  |  |  |  |  |
| Key results | 18 | Summarise key results with reference to study objectives | Discussion |  | Discussion |
| Limitations | 19 | Discuss limitations of the study, taking into account sources of potential bias or imprecision. Discuss both direction and magnitude of any potential bias | Discussion | RECORD 19.1: Discuss the implications of using data that were not created or collected to answer the specific research question(s). Include discussion of misclassification bias, unmeasured confounding, missing data, and changing eligibility over time, as they pertain to the study being reported. | Discussion |
| Interpretation | 20 | Give a cautious overall interpretation of results considering objectives, limitations, multiplicity of analyses, results from similar studies, and other relevant evidence | Discussion |  | Discussion |
| Generalisability | 21 | Discuss the generalisability (external validity) of the study results | N/A |  | N/A |
| **Other Information** |  |  |  |  |  |
| Funding | 22 | Give the source of funding and the role of the funders for the present study and, if applicable, for the original study on which the present article is based | Declarations |  | Declarations |
| Accessibility of protocol, raw data, and programming code |  | .. |  | RECORD 22.1: Authors should provide information on how to access any supplemental information such as the study protocol, raw data, or programming code. | Given the sensitive nature of the data, this information is not shared. |

## Appendix 3

**Eligible Clients:** Of the 881 129 adult clients in the Alliance EHR database in 2009-2019, 232,529 (26.4%) have ongoing primary care client indications, and 221,047 (25.1%) have at least one encounter in 2009-2019 (fully eligible).

**Table S1:**
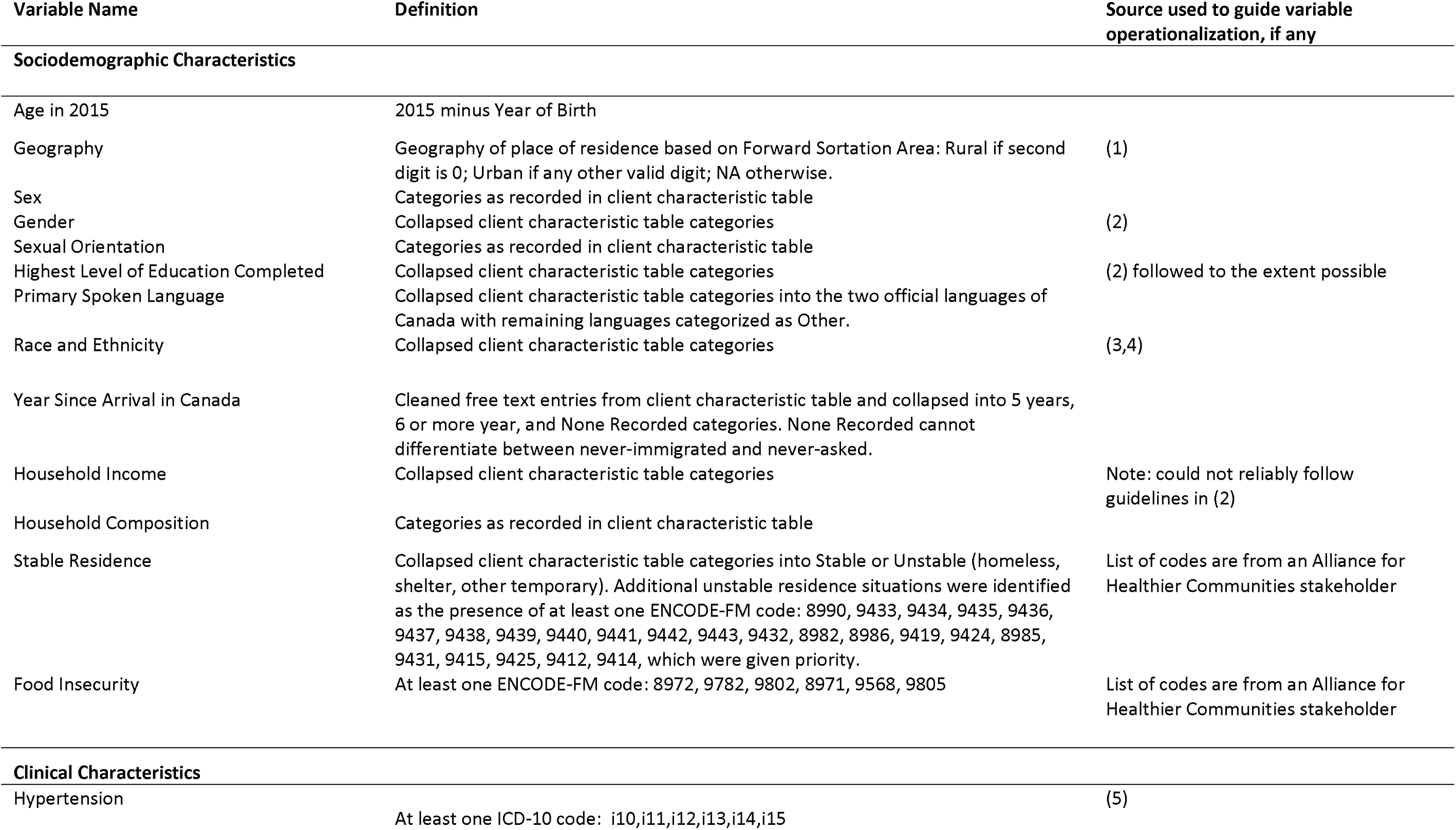
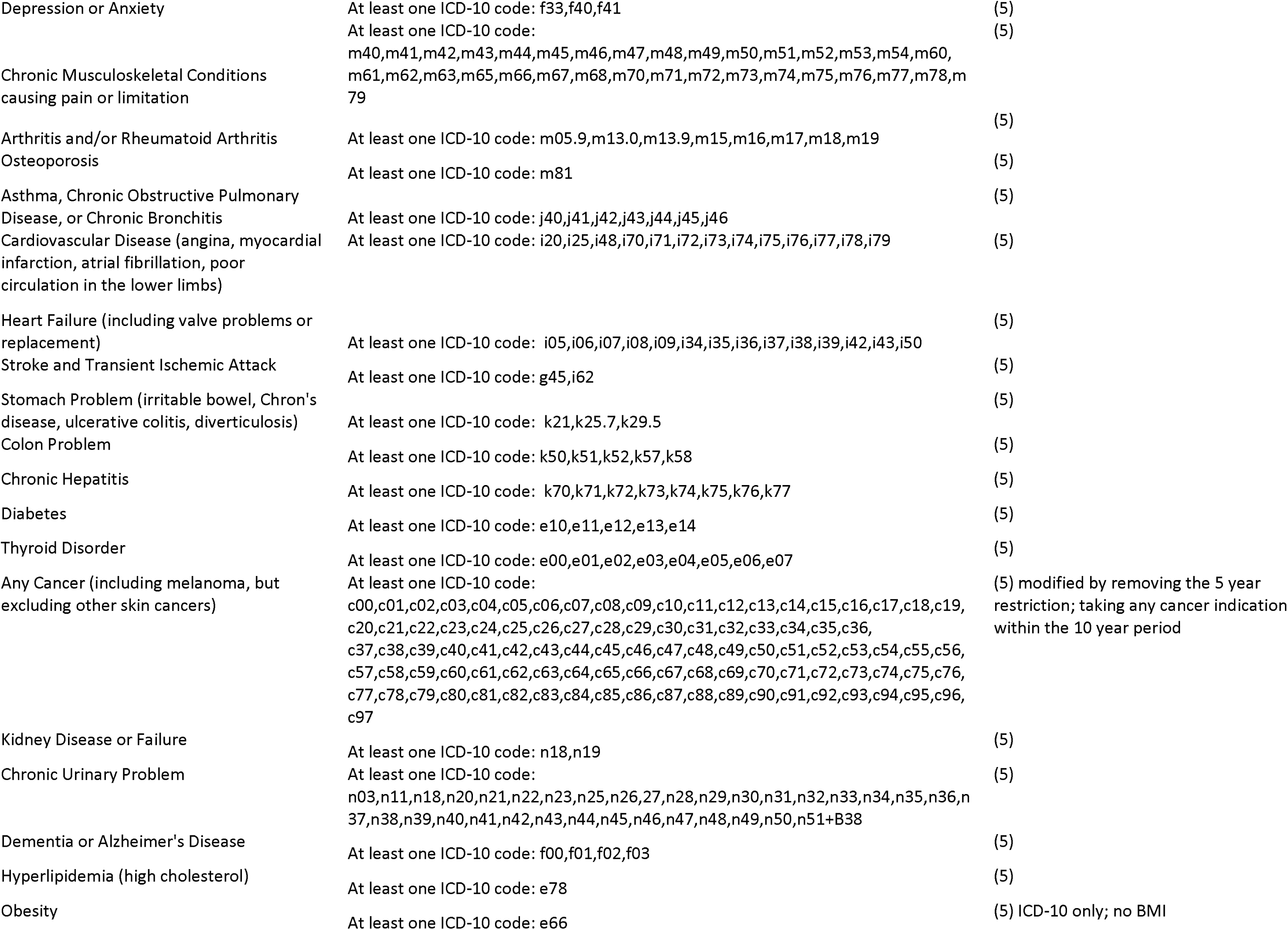
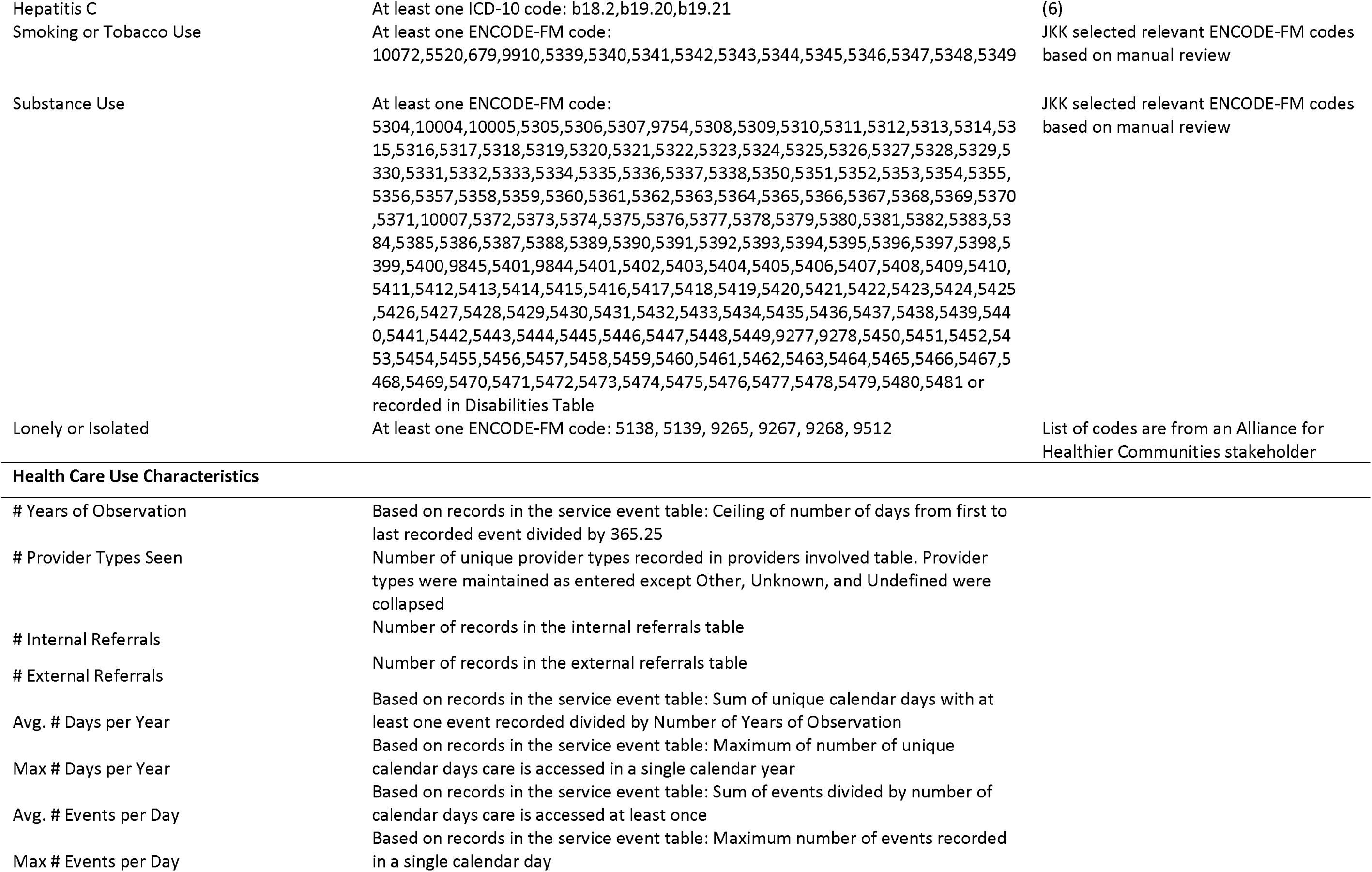
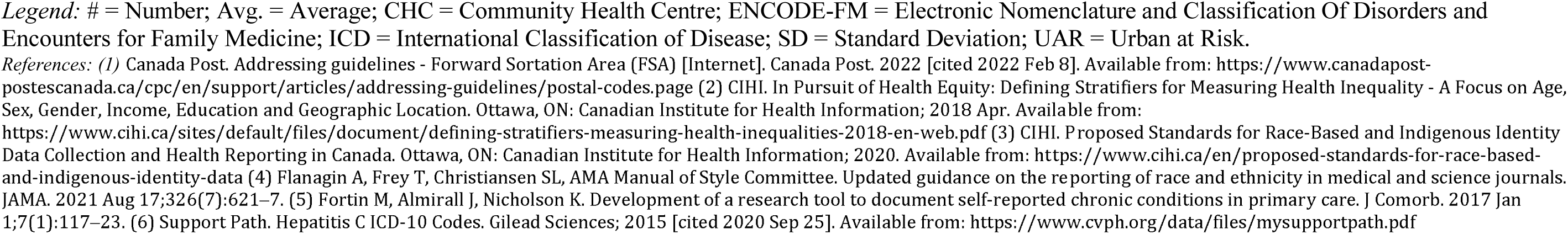
Characteristic variable definitions.

**Table S2:**
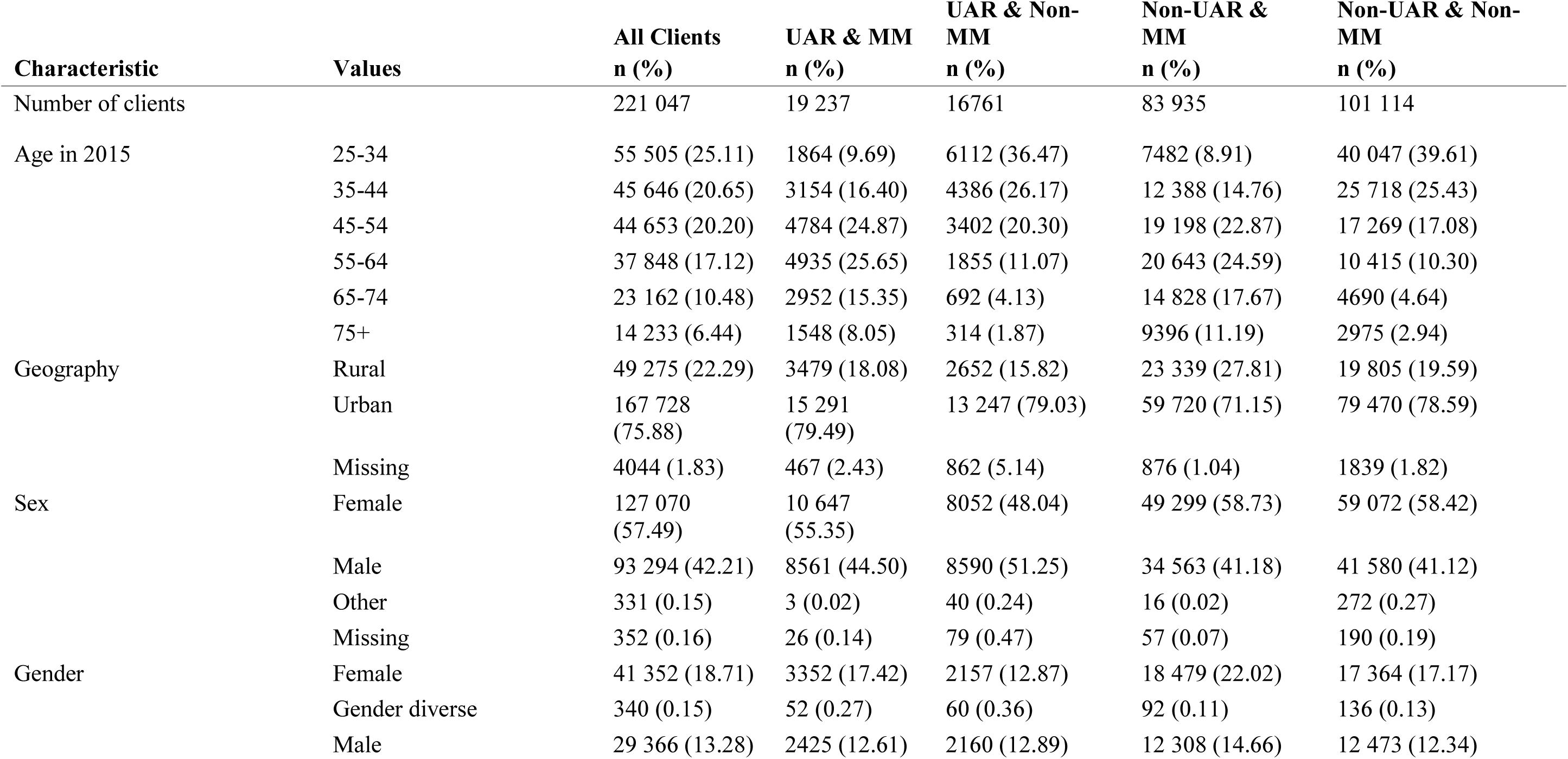
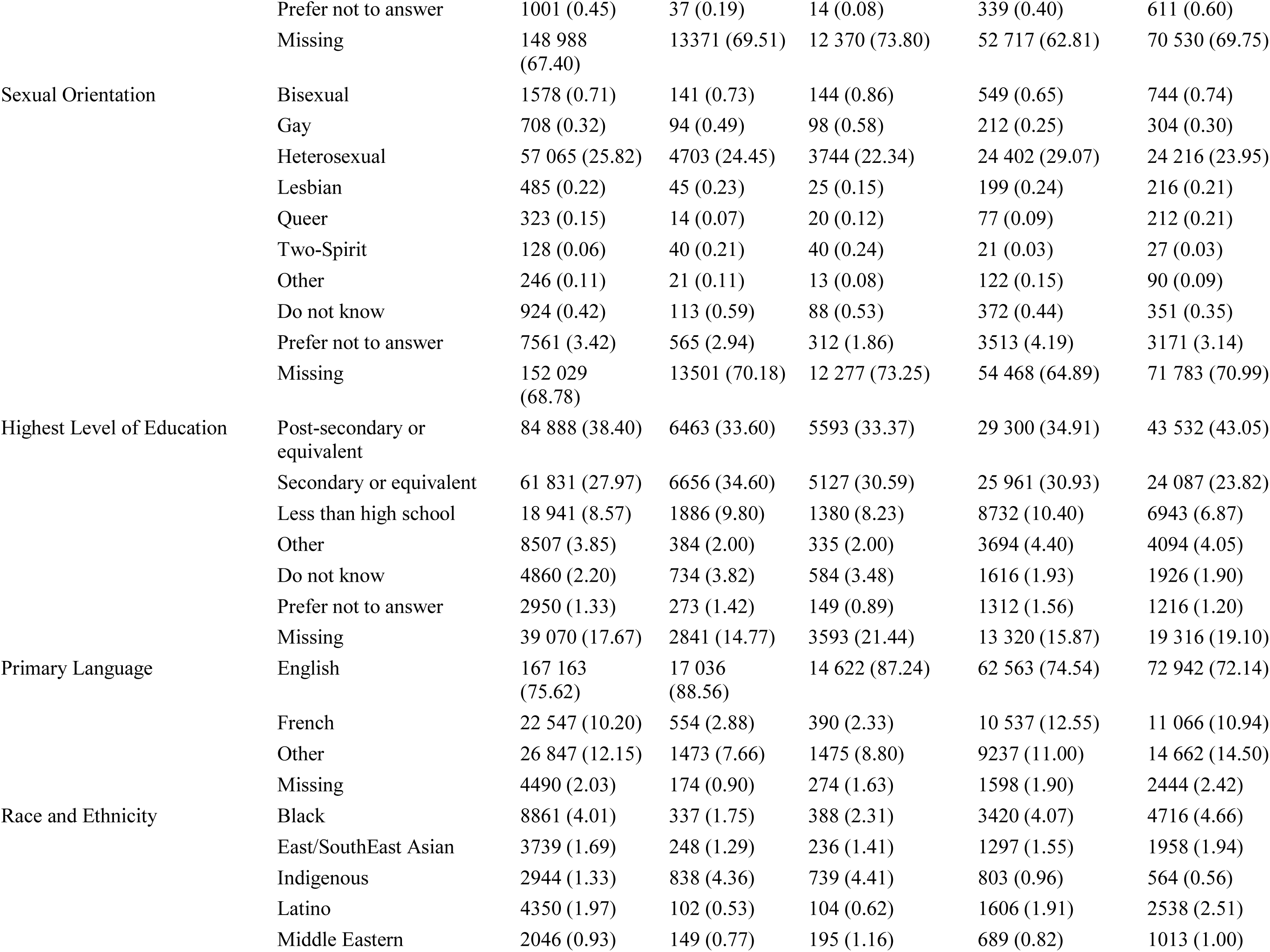
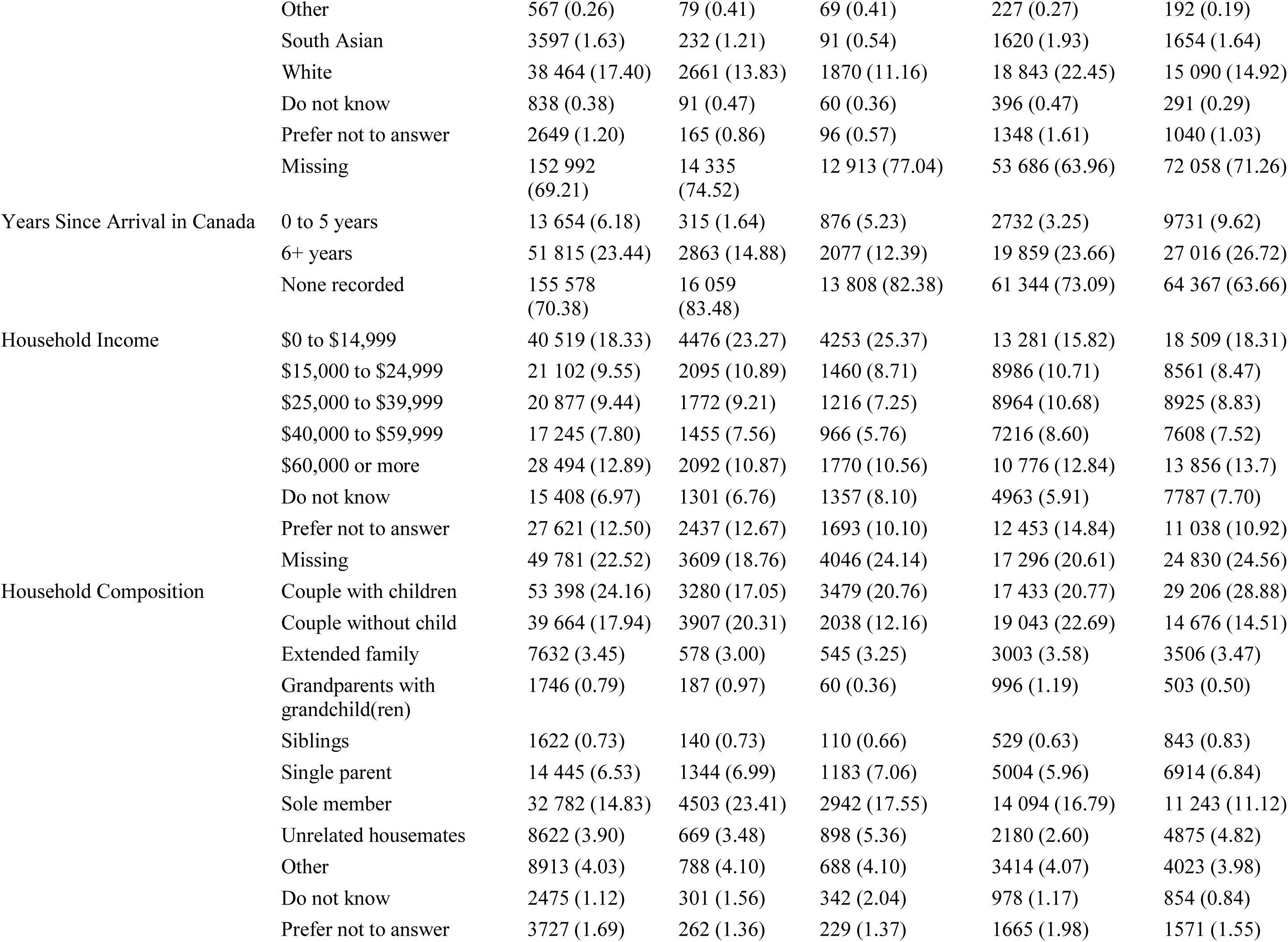
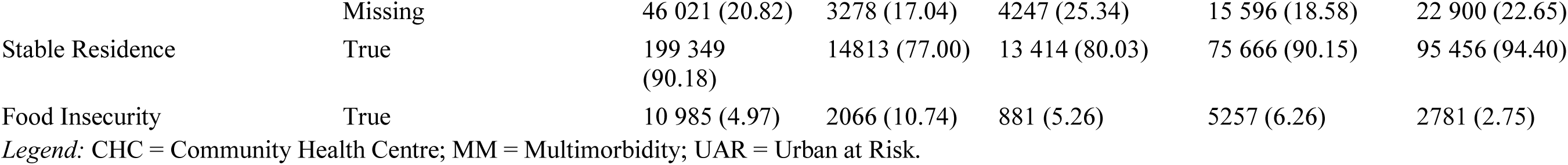
Sociodemographic characteristics with sub-strata.

**Table S3:** Health care use characteristics.

| Measure | Value | All Clients | UAR CHC | Rural CHC | Multimorbidity |
| --- | --- | --- | --- | --- | --- |
| # Years of Observation | Min, Median, Max | (1, 5, 11) | (1, 6, 11) | (1, 7, 11) | (1, 8, 11) |
|  | Mean (SD) | 5.6 (3.7) | 6.1 (3.8) | 6.7 (3.6) | 7.4 (3.3) |
| 11 Years of Observation | n (%) | 36 724 (16.6) | 7976 (22.2) | 5374 (25) | 29 062 (28.2) |
| # Provider Types Seen | Min, Median, Max | (0, 4, 19) | (0, 5, 19) | (0, 5, 14) | (0, 6, 19) |
|  | Mean (SD) | 4.5 (2.3) | 5.1 (2.6) | 4.8 (2.1) | 5.8 (2.2) |
| # Internal Referrals | Min, Median, Max | (0, 0, 300) | (0, 1, 300) | (0, 0, 51) | (0, 1, 300) |
|  | Mean (SD) | 1.7 (4.3) | 2.8 (7.0) | 1.4 (2.8) | 2.8 (5.7) |
| # External Referrals | Min, Median, Max | (0, 1, 309) | (0, 2, 309) | (0, 1, 46) | (0, 3, 309) |
|  | Mean (SD) | 2.9 (4.5) | 3.8 (5.9) | 2.5 (3.2) | 4.8 (5.5) |
| Avg. # Days/Year | Min, Median, Max | (0.2, 6, 176.9) | (0.2, 6.9, 129.7) | (0.2, 6.2, 120.3) | (0.3, 9.2, 176.9) |
|  | Mean (SD) | 8 (7.4) | 9.4 (8.9) | 8 (6.7) | 11.4 (8.4) |
| Max # Days/Year | Min, Median, Max | (1, 10, 349) | (1, 12, 245) | (1, 11, 349) | (1, 17, 349) |
|  | Mean (SD) | 13.7 (13) | 16.8 (16.6) | 14.2 (12.1) | 20.3 (14.4) |
| Avg. # Events/Day | Min, Median, Max | (1, 1.2, 66) | (1, 1.2, 29) | (1, 1.2, 31) | (1, 1.2, 31) |
|  | Mean (SD) | 1.3 (0.5) | 1.3 (0.3) | 1.3 (0.3) | 1.3 (0.3) |
| Max # Events/Day | Min, Median, Max | (1, 3, 635) | (1, 3, 635) | (1, 3, 224) | (1, 3, 635) |
|  | Mean (SD) | 3.9 (7.8) | 4.1 (8.2) | 3.8 (6.1) | 5.5 (10.7) |

*Legend: # = Number; Avg. = Average; CHC = Community Health Centre; SD = Standard Deviation; UAR = Urban at Risk.*

**Table S4:**
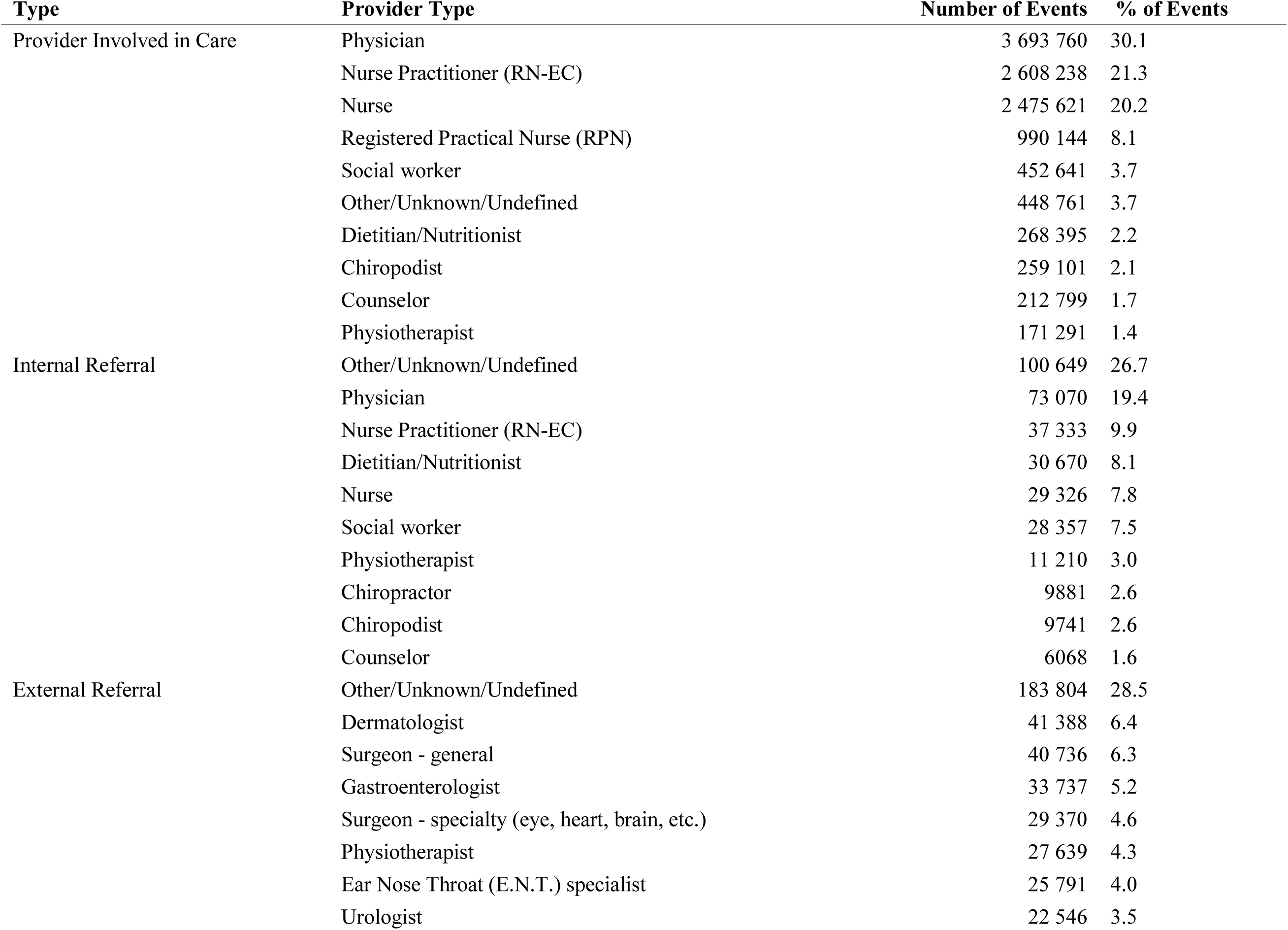
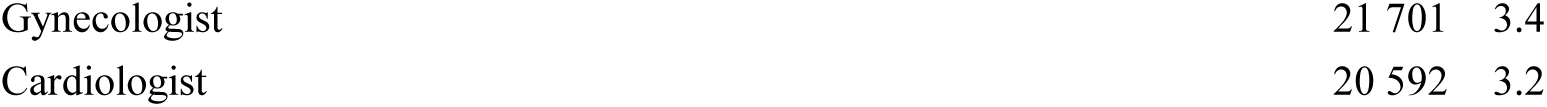
Provider type counts.

**Table S5:**
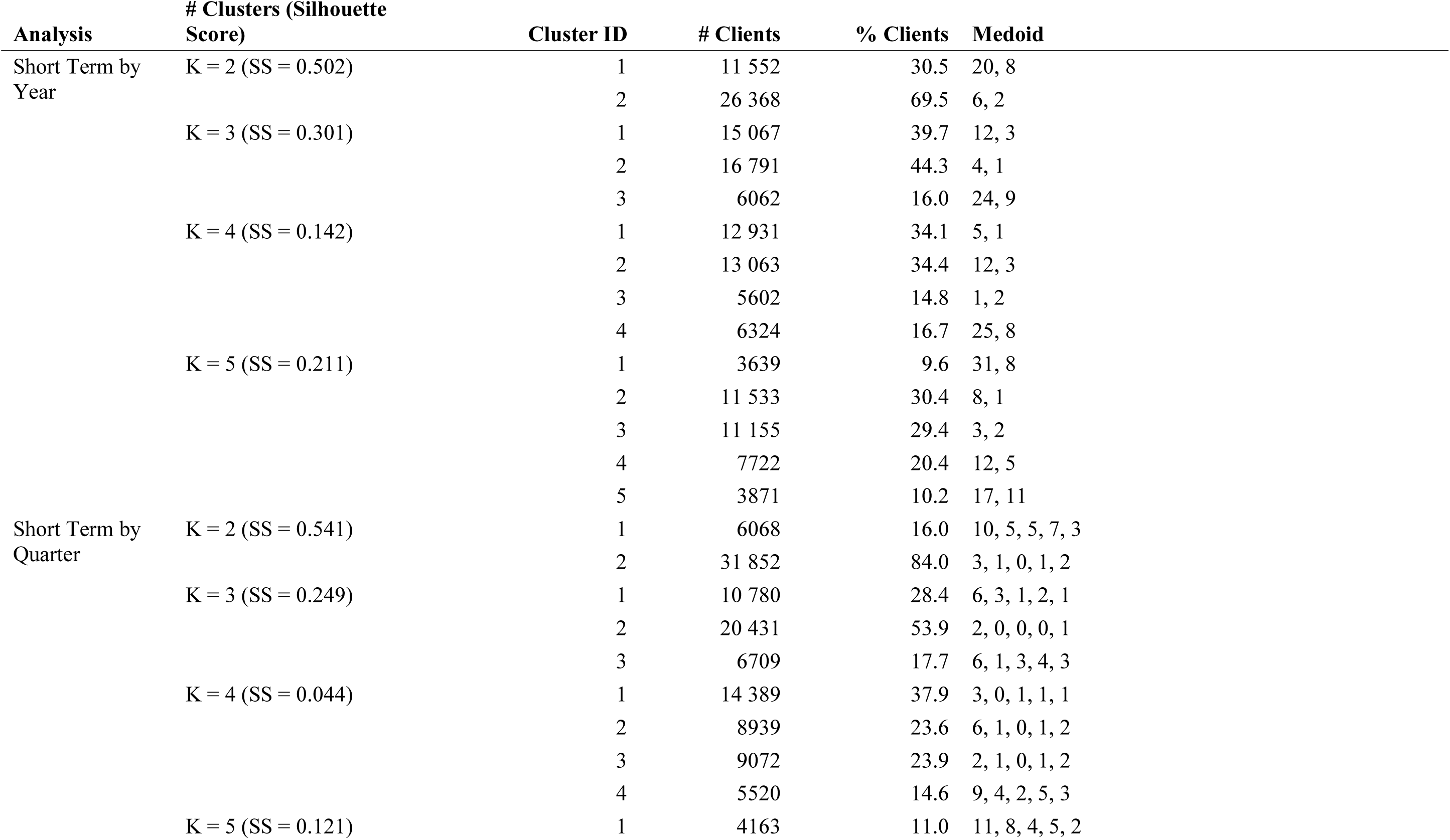
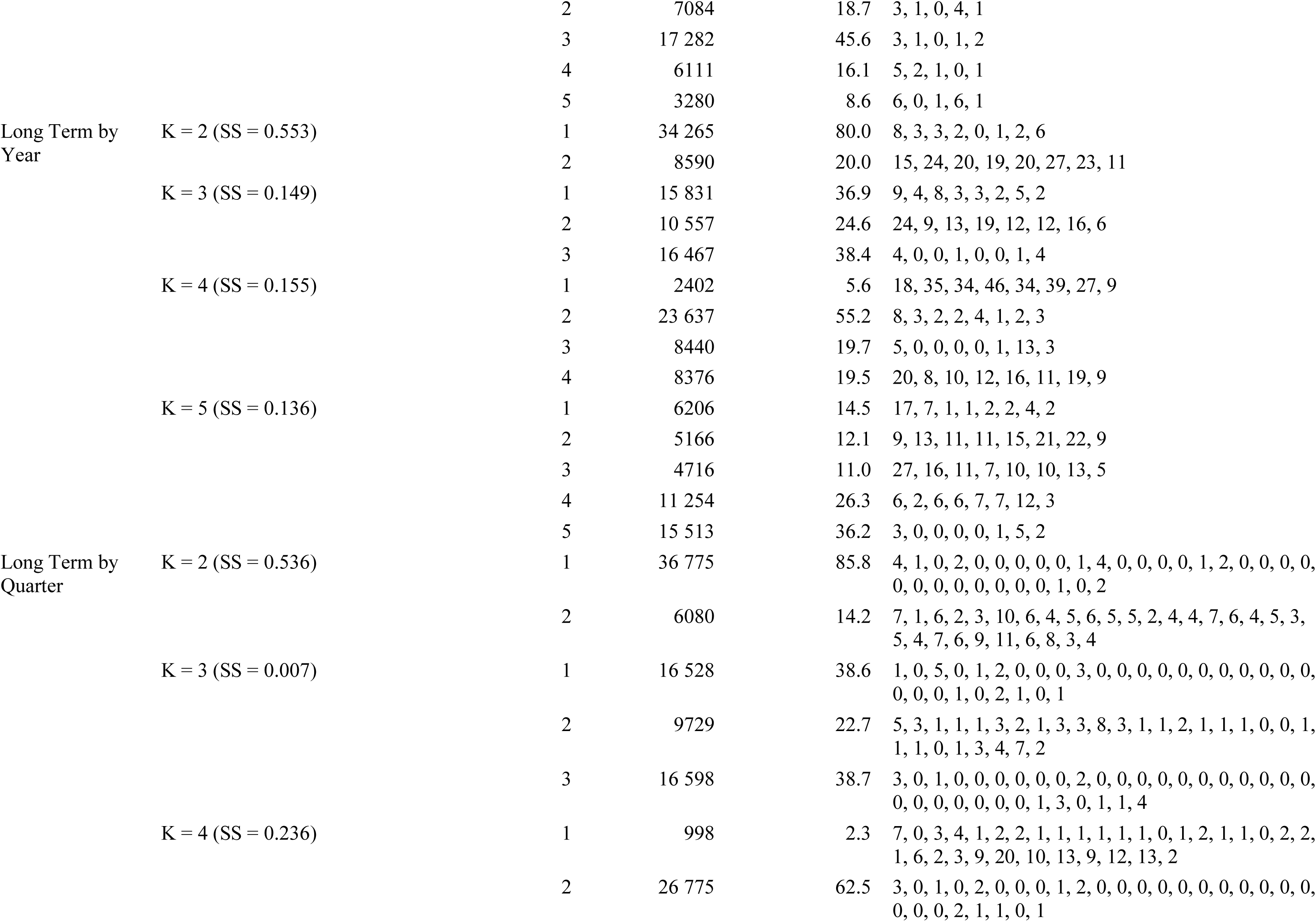
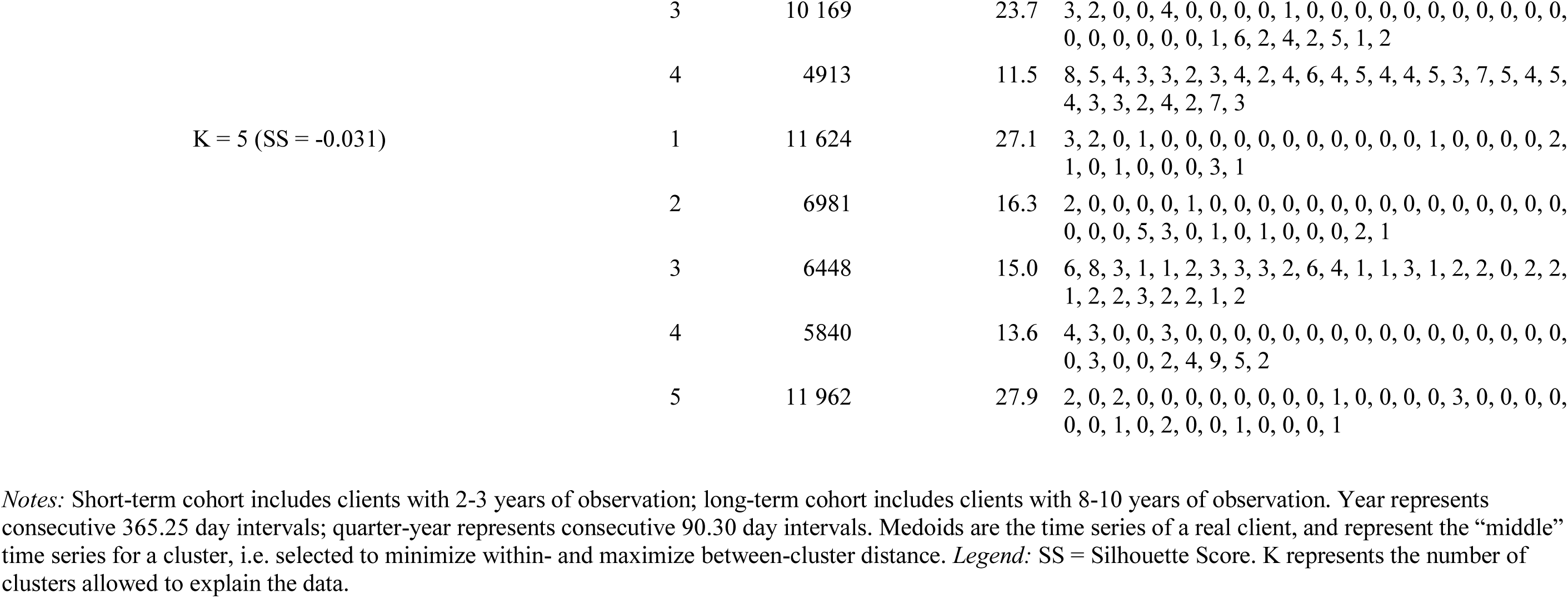
Time series clustering of care access frequency.

### Eleven-year period prevalence technical details

Since not all clients receive care from CHCs 2009-2019, they are not all at-risk of condition indications in their electronic health record for the entire calendar-based period of observation. Thus, the denominator requires estimation of the average or mid-point size of the population. This is challenging given that primary care electronic health records represent an open cohort with no standard expectation for frequency of care, and the overall number of clients receiving care increases across calendar time (see Supplementary Figure 1). We used the following process to calculate 11-year period prevalence:

Numerator: number of clients with at least one relevant code at any point from 2009 through 2019.

Denominator: First, we calculated the median number of calendar-based years of observation across all eligible clients (i.e., median number of “at-risk” years): 5 years. Second, we calculated the number of clients who received any type of care at least once in each of the seven possible five-year intervals (2009-13; 2010-14; 2011-15; 2012-16; 2013-17; 2014-18; 2015-19), representing the size of the population within each of those five-year intervals. Finally, the median size of those seven cohorts was used as the denominator, representing the overall average size of the population across 11 years.

The same process was followed to get estimates for the entire eligible population and for the subset of clients who receive care from urban at risk community health centres.

